# COVID-19 convalescent plasma and randomized clinical trials: explaining conflicting outcomes and finding signals of efficacy

**DOI:** 10.1101/2021.09.07.21263194

**Authors:** Daniele Focosi, Massimo Franchini, Liise-anne Pirofski, Thierry Burnouf, Nigel Paneth, Michael J. Joyner, Arturo Casadevall

## Abstract

Convalescent plasma (CP) recurs as a frontline treatment in epidemics because it is available as soon as there are survivors. The COVID-19 pandemic represented the first large-scale opportunity to shed light into mechanisms of action, safety and efficacy of CP using modern evidence-based medicine approaches. Studies ranging from observational case series to randomized controlled trials (RCT) have reported highly variable efficacy results for COVID-19 CP (CCP), resulting in uncertainty. Reasons for CCP success and failure may be hidden in study details, which are usually difficult to explain to physicians and the public but provide fertile ground for designing next-generation studies. We analyzed variables associated with efficacy such as clinical settings, disease severity, CCP SARS-CoV-2 antibody levels and function, dose, timing of administration (variously defined as time from onset of symptoms, molecular diagnosis, diagnosis of pneumonia, or hospitalization, or by serostatus), outcomes (defined as hospitalization, requirement for ventilation, clinical improvement or mortality), CCP provenance and time for collection, and criteria for efficacy. Focusing only on the results from the 30 available RCTs we noted that these were more likely to show signals of efficacy, including reductions in mortality, if the plasma neutralizing titer was ≥ 160 and the time to randomization was ≤ 9 days, consistent with passive antibody therapy efficacy requiring dosing with sufficient antibody. The fact that most studies revealed signals of efficacy despite variability in CCP and its use suggest likely therapeutic effects that become apparent despite the data noise. Despite the recent WHO guidelines discouraging CCP usage, the Omicron variant of concern is reminding us the superiority of polyclonal antibody therapies over monoclonal antibodies, and CCP from vaccinated convalescents is likely to be evaluated soon

## Introduction

In the first 21 years of the 21^st^ century humanity has experienced six major multinational epidemics. The agents involved were SARS-CoV, MERS-CoV, influenza A(H_1_N_1_), Ebola, Zika and SARS-CoV-2 viruses. For the five most lethal of these outbreaks the response included the use of convalescent plasma (CP) (reviewed in (1, 2)) and it was considered for the less lethal sixth (Zika virus). The attraction of CP is that it is readily available as soon as there are convalescing survivors, that unlike drugs or monoclonal antibodies it needs no development, and it is polyclonal, affordable and deployable even in resource poor countries. Despite suffering from some logistical hurdles (dedicated collection, testing and handling procedures, heterogeneity, standardization of the therapeutic dose, blood type matching, and intravenous delivery), CP has been proposed as a first line response to new pandemics (3) and was deployed during the COVID-19 pandemic in March 2020 in countries that experienced the early waves of disease such as China (4, 5) and Italy (6). The relatively low COVID- 19 case-fatality rate (compared to the other epidemic agents noted above) allowed for testing of CP across a wider spectrum of disease severity.

While in early 2020 most clinical use was reported in case series or small phase II clinical trials (7), beginning in late March 2020 the US expanded access program (EAP) generated a large and robust treatment dataset, with insights on safety and optimal use. This database provided the first clear evidence that CP is safe, which was important given that early in the pandemic there were significant concerns about antibody-dependent enhancement (8). Later, an analysis of the first 3082 patents within the EAP database provided evidence that associated early administration of high titer CCP to non-ventilated hospitalized patients with reduced mortality (9). Before the FDA granted emergency use authorization (EUA), the US EAP provided CCP to as many as 94,287 patients.

During the past year, many studies employing either randomized controls (RCT) or propensity score- matched (PSM) controls have been published. RCTs and PSM studies reported so far have had largely opposite outcomes, with most but not all RCTs finding little overall effect on mortality while the PSM and many smaller trials reporting mortality benefits. Several RCTs did not have mortality as a primary endpoint or it was part of a composite endpoint (5, 10–12). These disparate results have led to confusion for both the public and the clinicians, leading to reduced enthusiasm for the use of CP, in part because RCT data is more influential in affecting the opinion of many physicians, specialty societies and government regulators.

As with any other medical treatment, several key factors should be taken into account when evaluating a trial, including the indication (which can be estimated by timing or clinical severity), the therapeutic dose and the intended outcomes. The choices made by the trial designers determine whether the trial will demonstrate clinical benefit. While much attention is appropriately focused on the performance features of clinical trials (sample size, fidelity to randomization, appropriate analysis), the biological rationale for the hypothesis being tested is critically important but not always taken into account.

## Methods

On December 13, 2021, we searched PubMed (which is also indexing the medRxiv prepublishing server) for clinical trials of CCP in COVID19, focusing on RCTs and PSM studies only. Each study was analyzed for the following variables: NCT identifier, recruitment, randomization strategy, type of control arm, baseline patient status, median neutralizing antibody (nAb) titer in both recipients (before CCP transfusion) and CCP units, type of viral neutralization test (VNT), primary endpoint, signals of efficacy, and reasons for failure At the same date, the ClinicalTrials.gov database was searched for CCP RCTs worldwide having as status “completed”, “active, not yet recruiting” or “recruiting”.

## Results

PubMed search retrieved 28 RCTs and 13 PSM studies about CCP, whose main variables are summarized in Tables 2 and 3. The characteristics of the VNTs used are summarized in Table 1. The variables were reconciled in 4 major topics, discussed in the following sections: the indication, the therapeutic doses, the relevance of CCP to the viral variant, and the intended outcome.

**Table 1.**
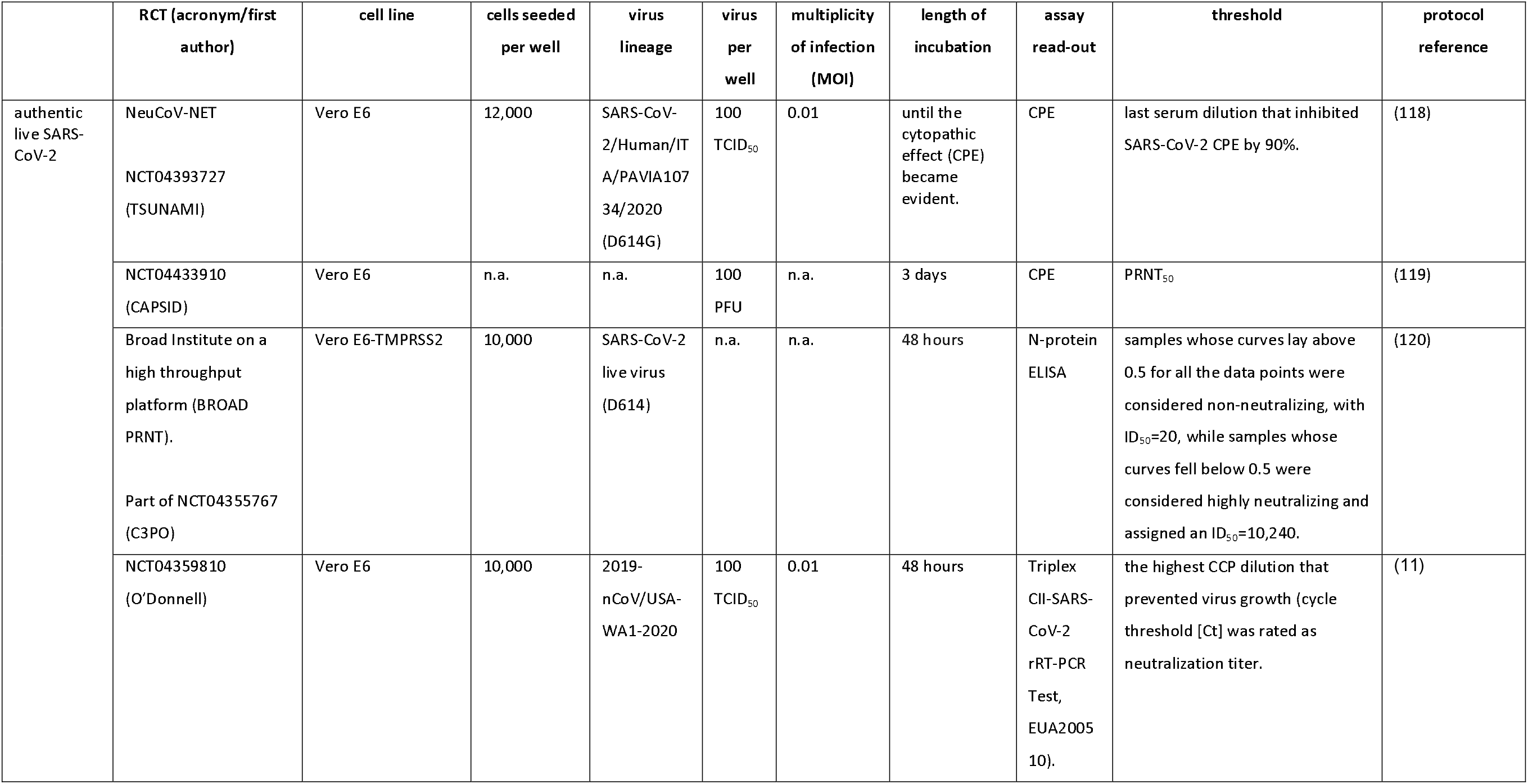
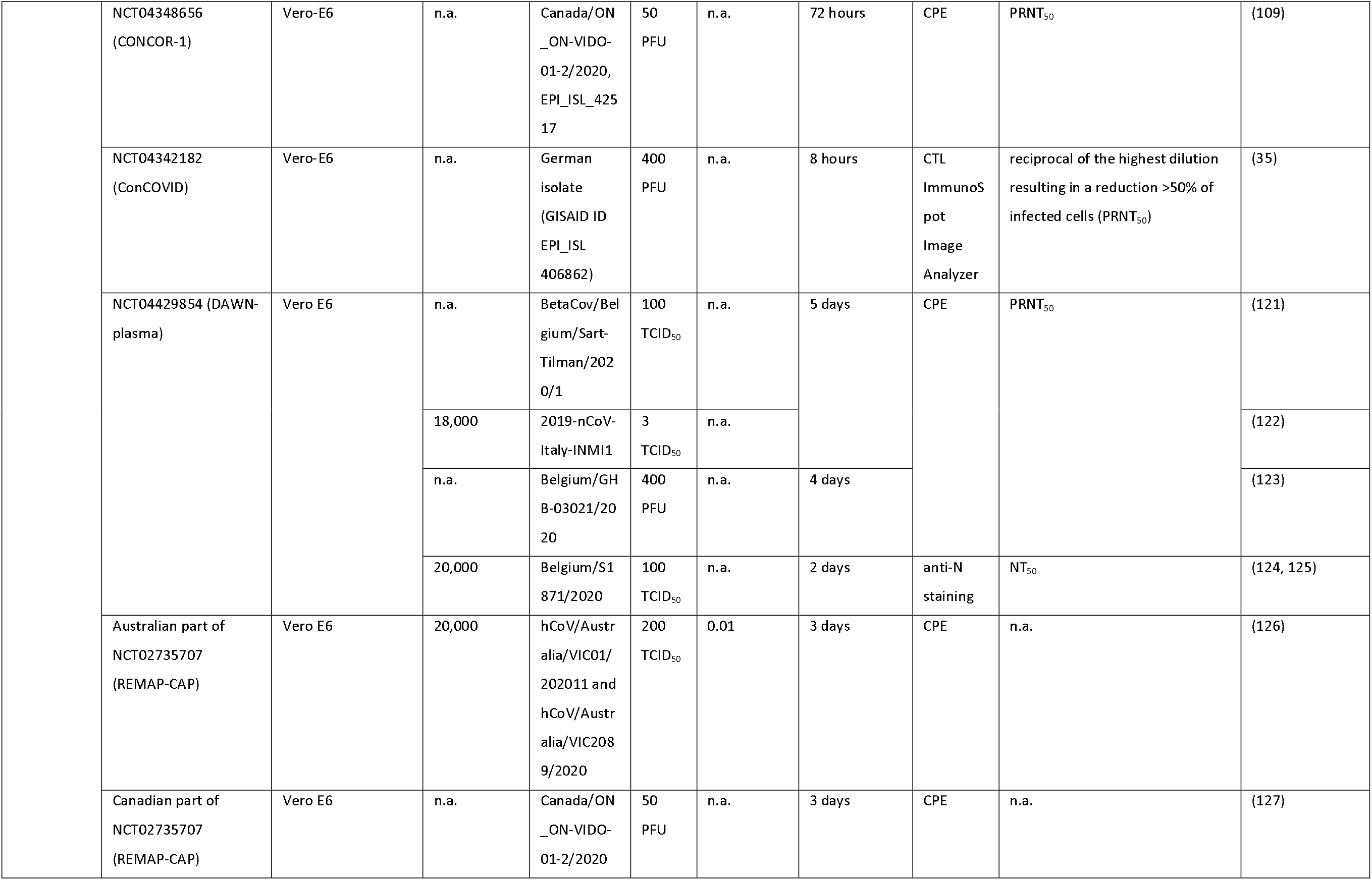
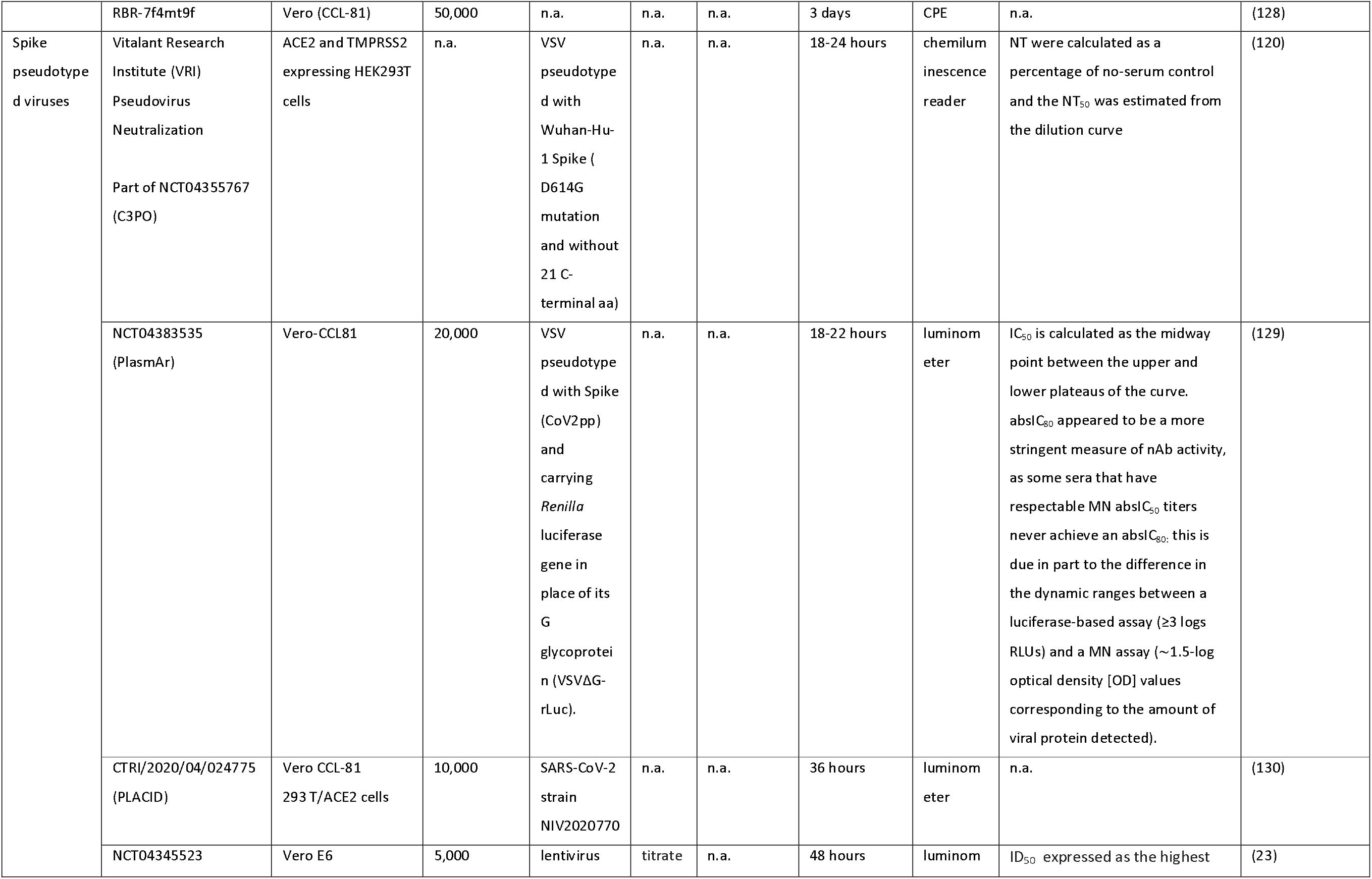
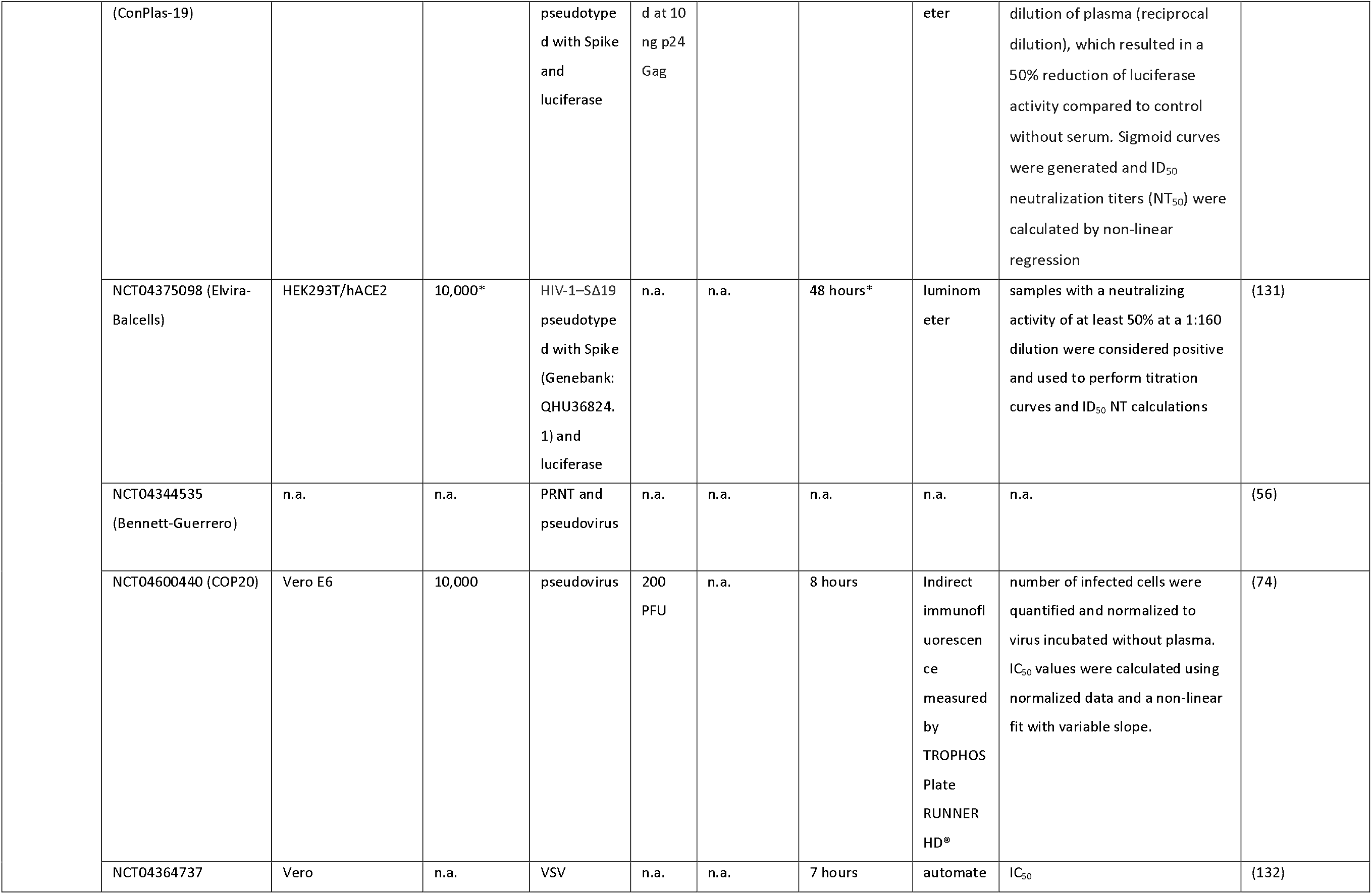
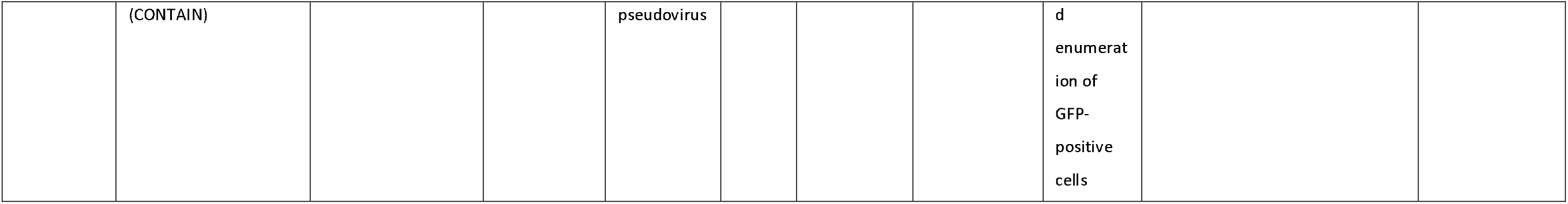
Details of viral neutralization tests (VNT) employed in CCP RCTs. Information was retrieved from original article (including Supplementary Appendix). Whenever not reported, the corresponding author was contacted (marked with *). If the details could not be retrieved the field is labelled “n.a.” (not available). IC: inhibitory concentration. NT: neutralization titer. PFU: plaque-forming unit; PRNT : plaque reduction neutralization test.

**Table 2.**
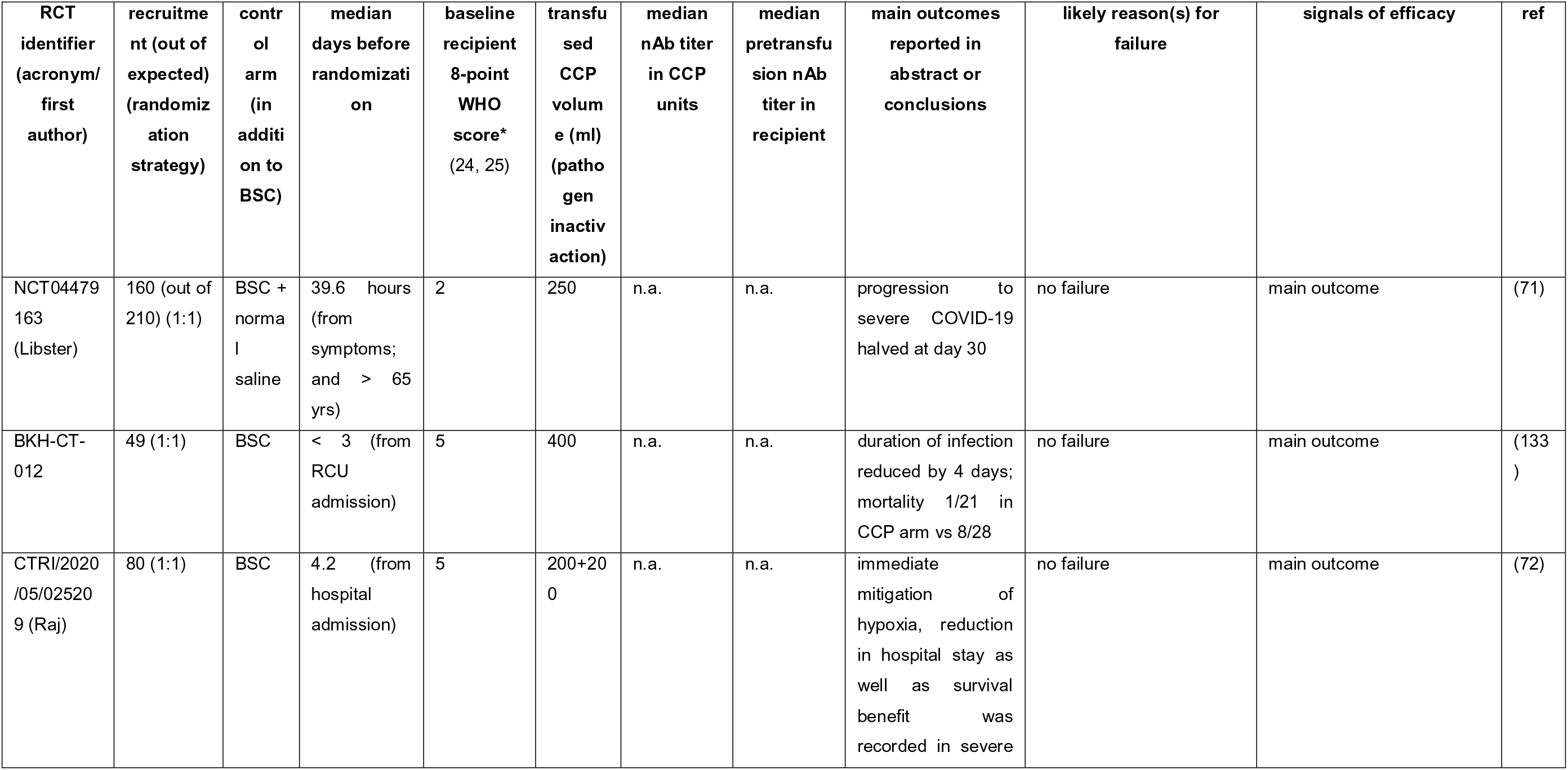
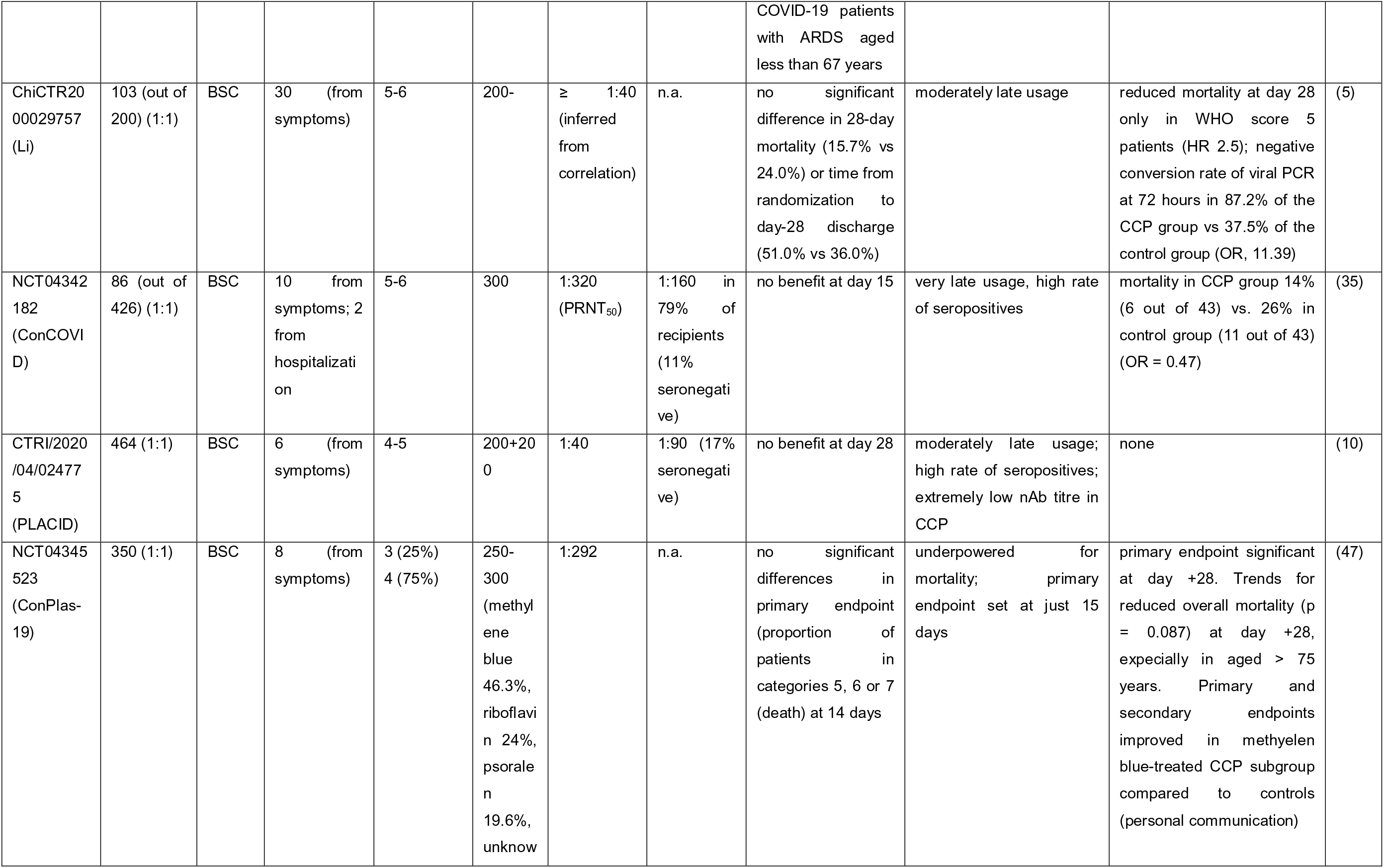
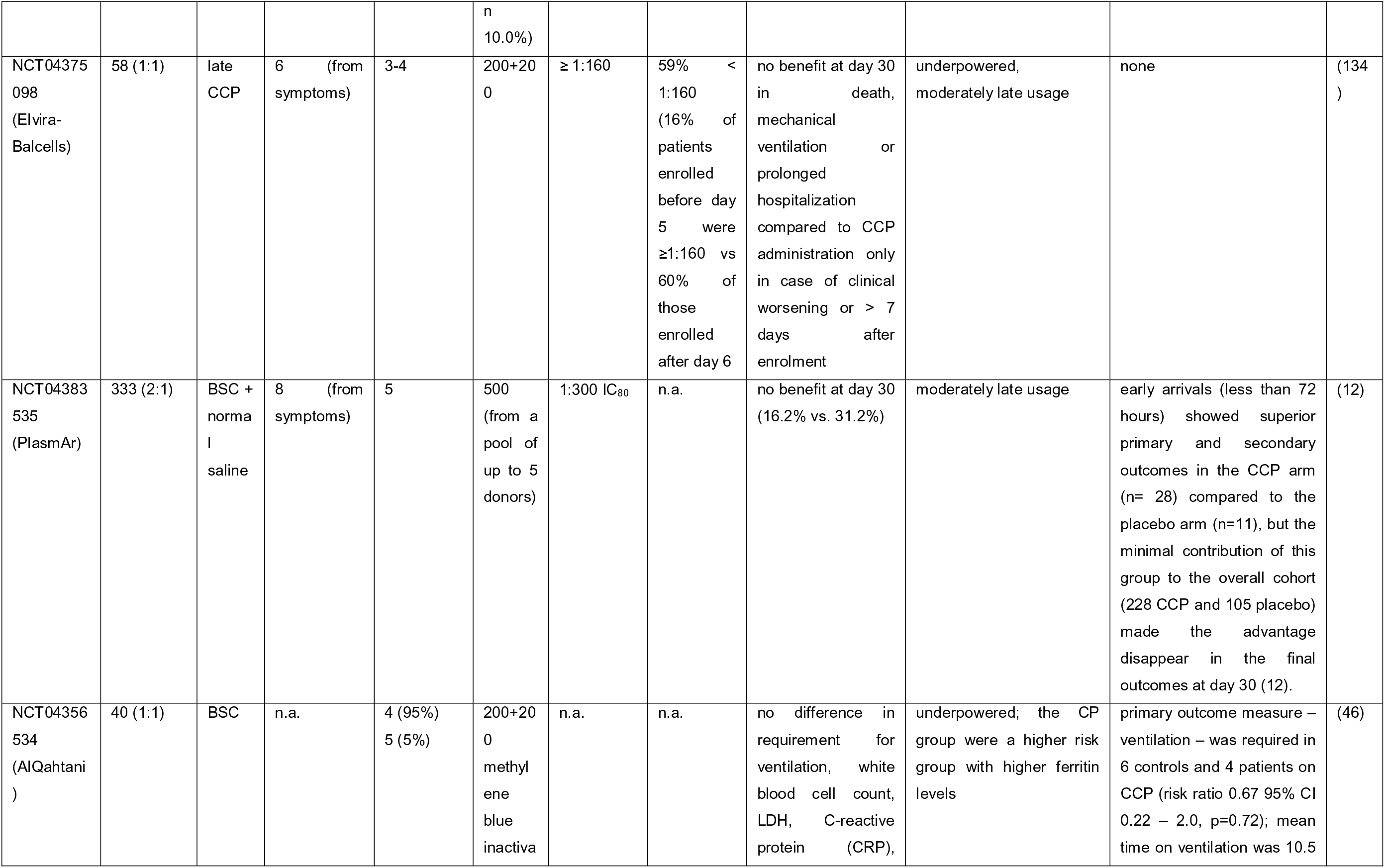
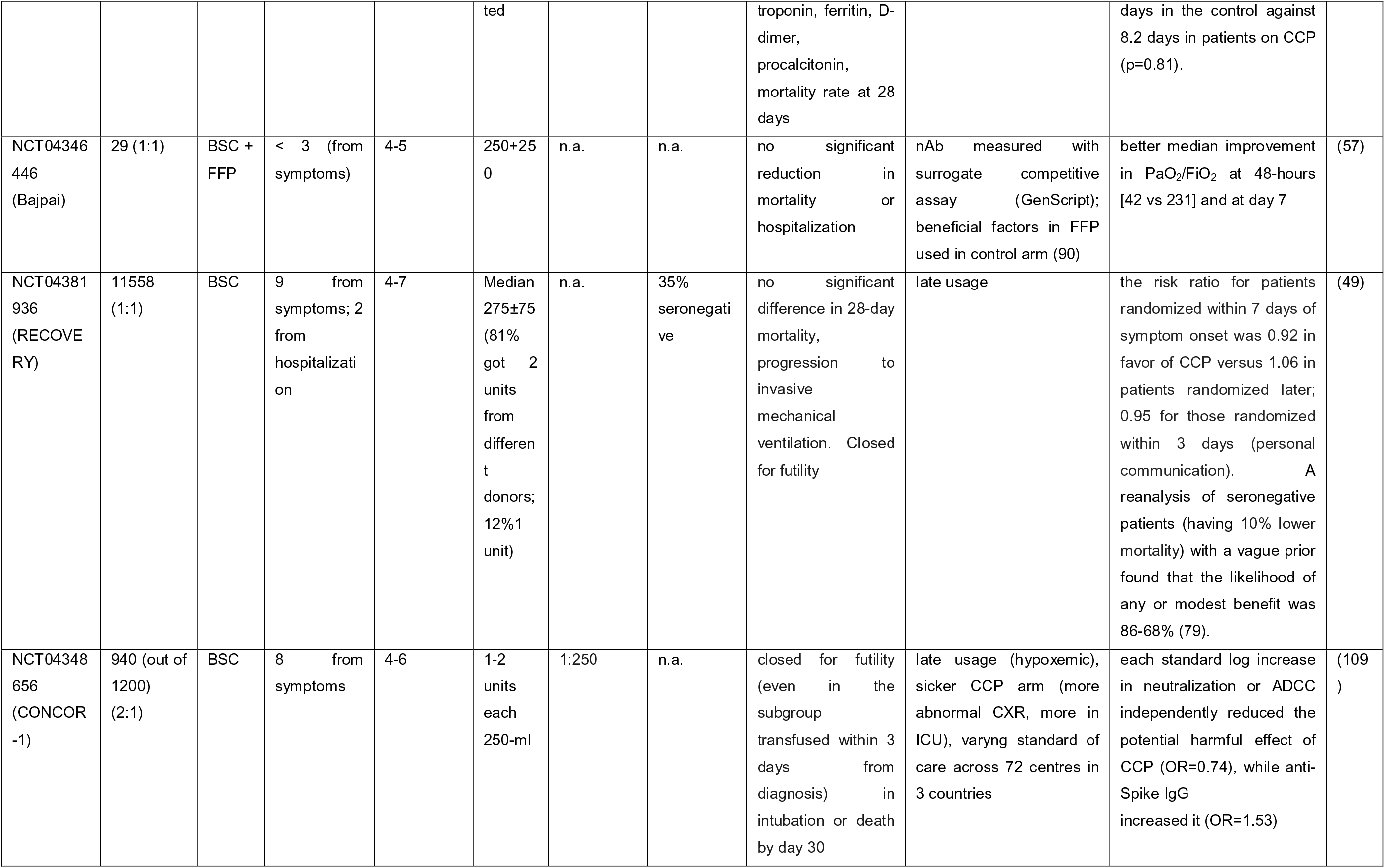
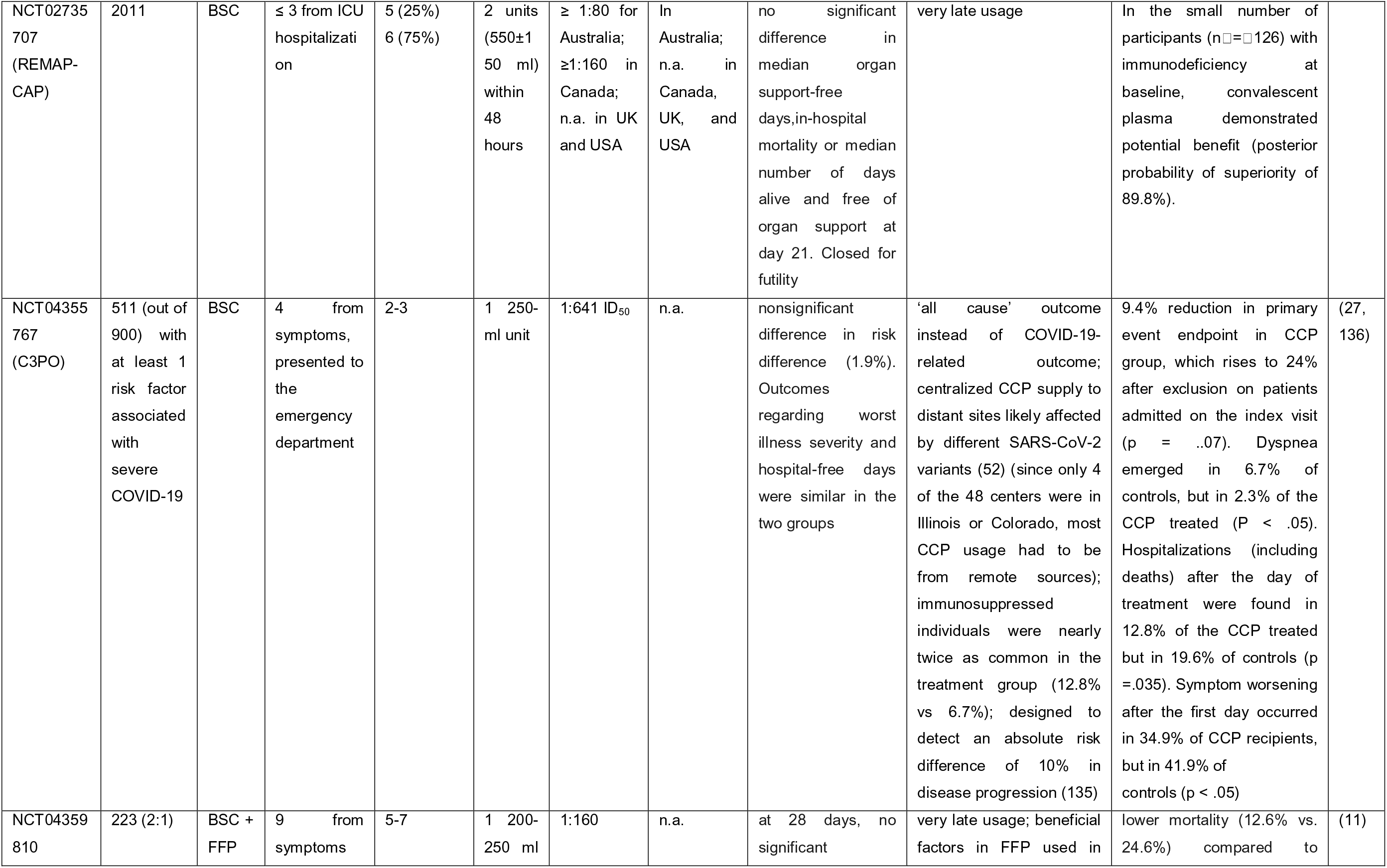
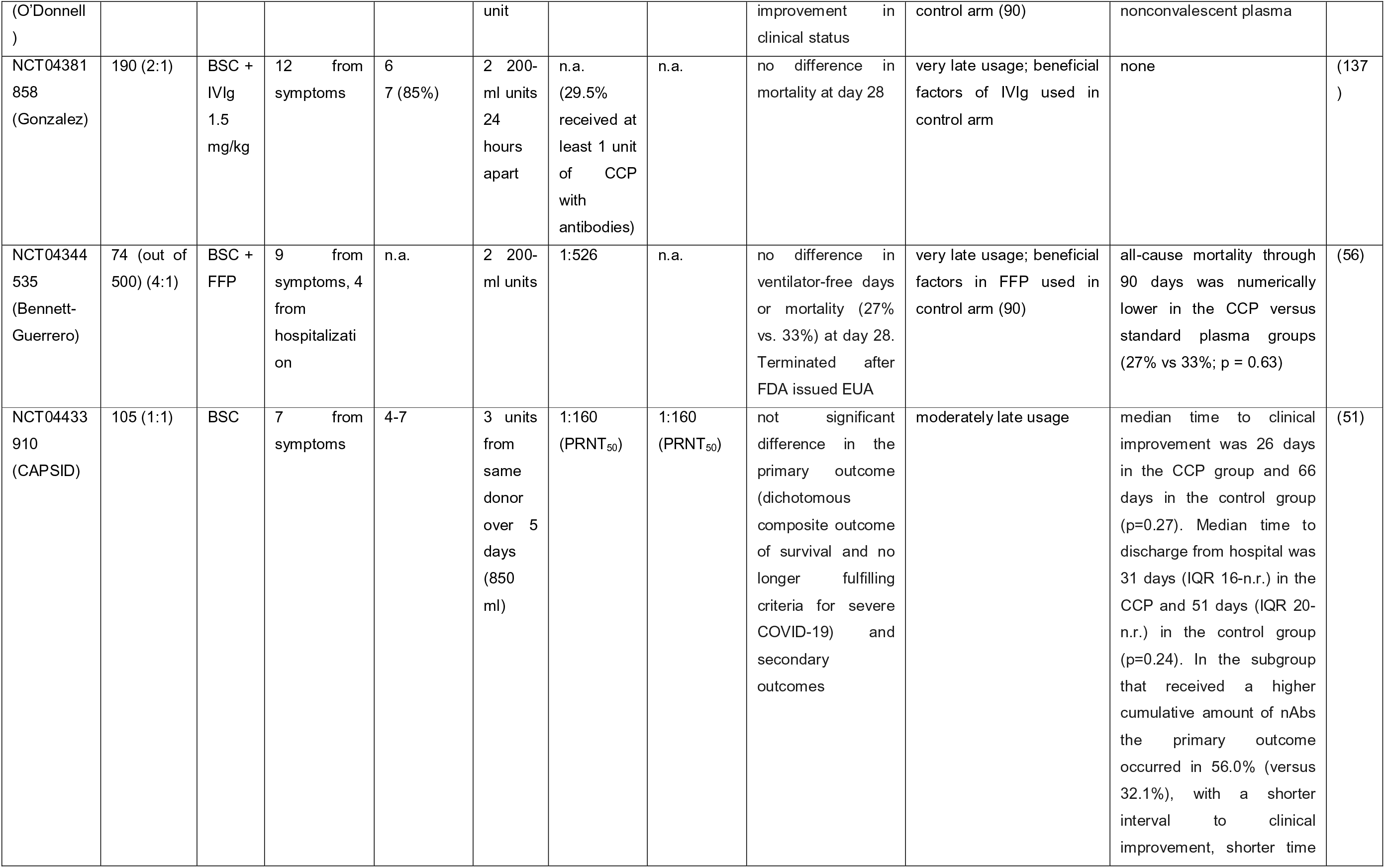
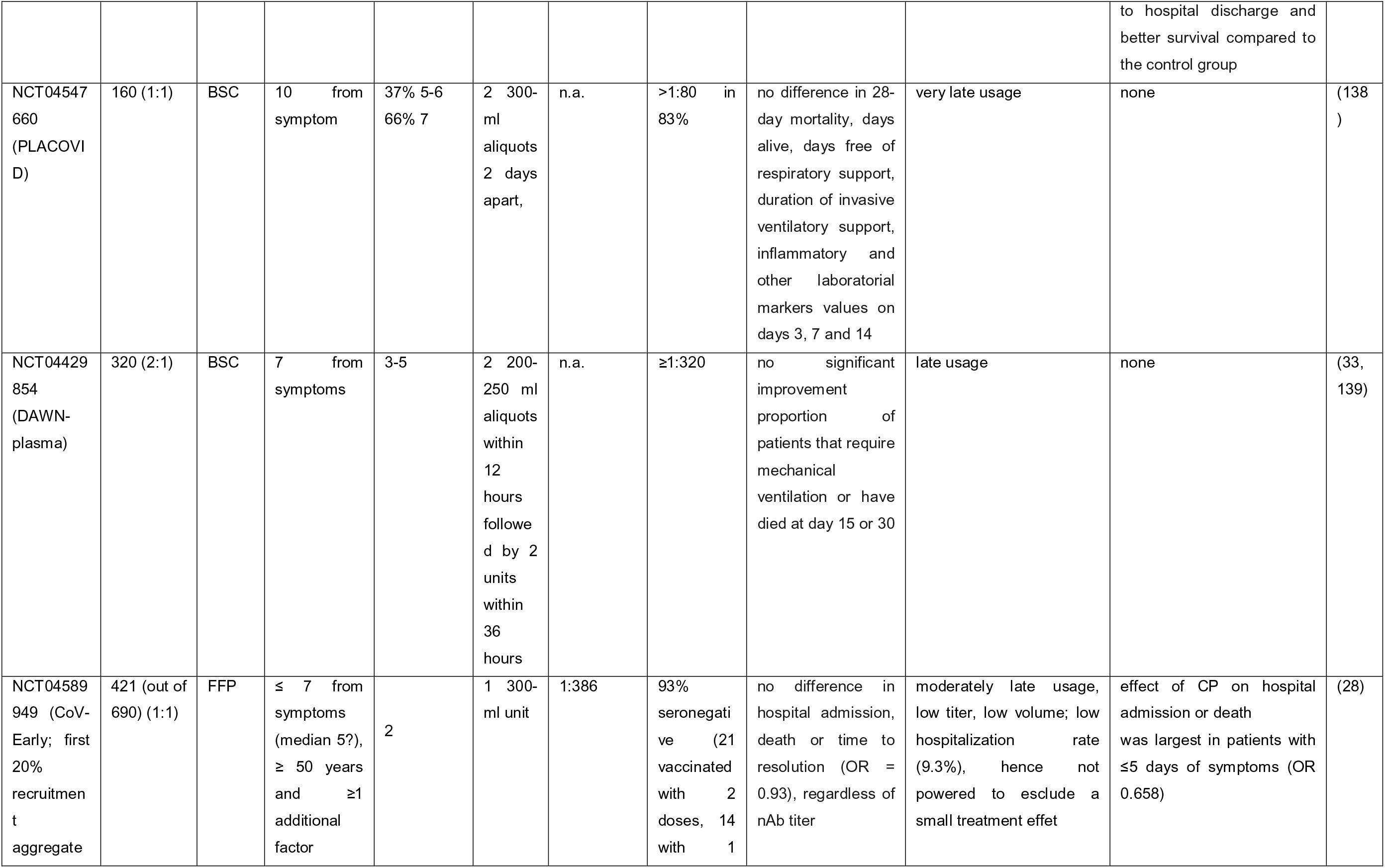
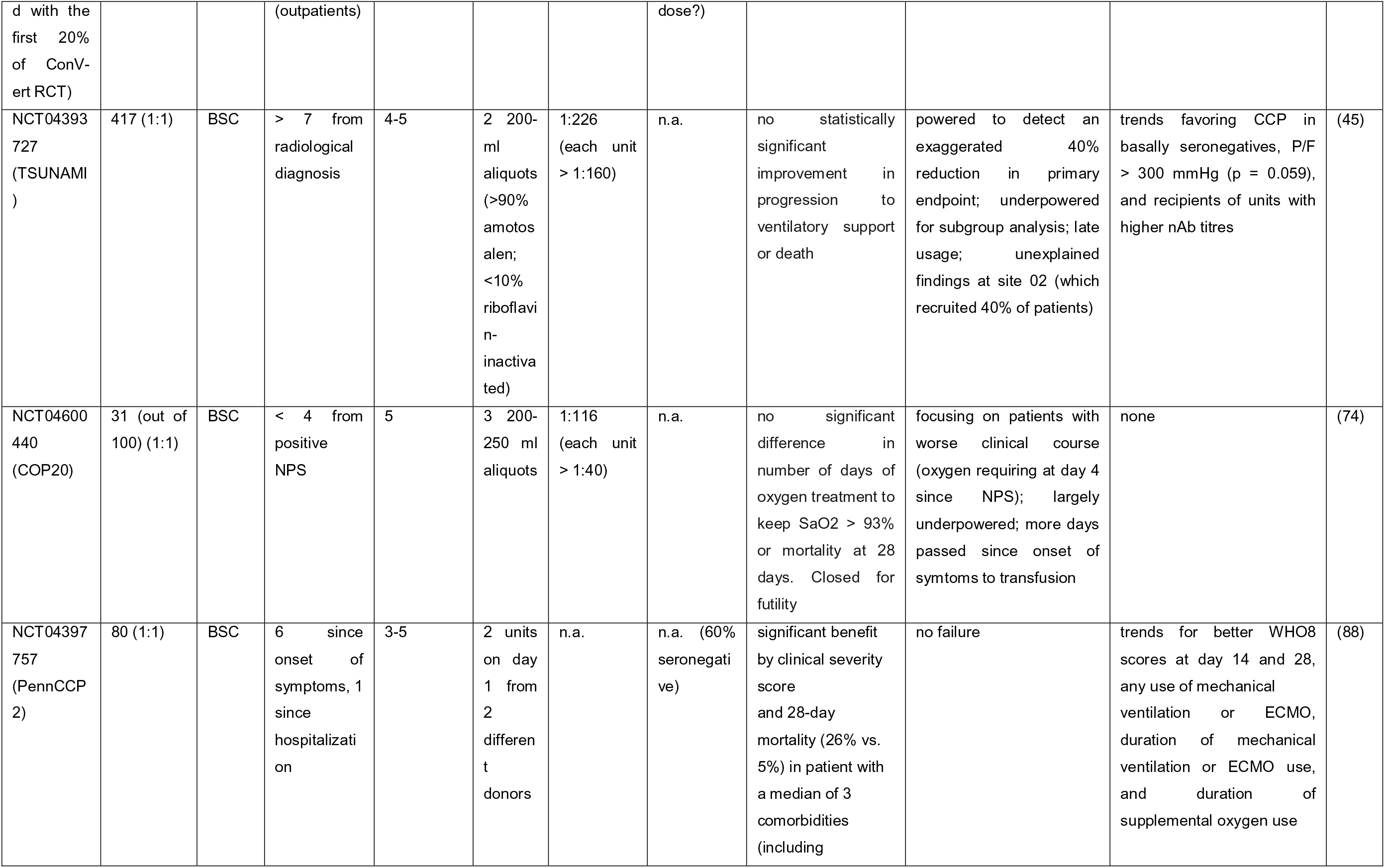
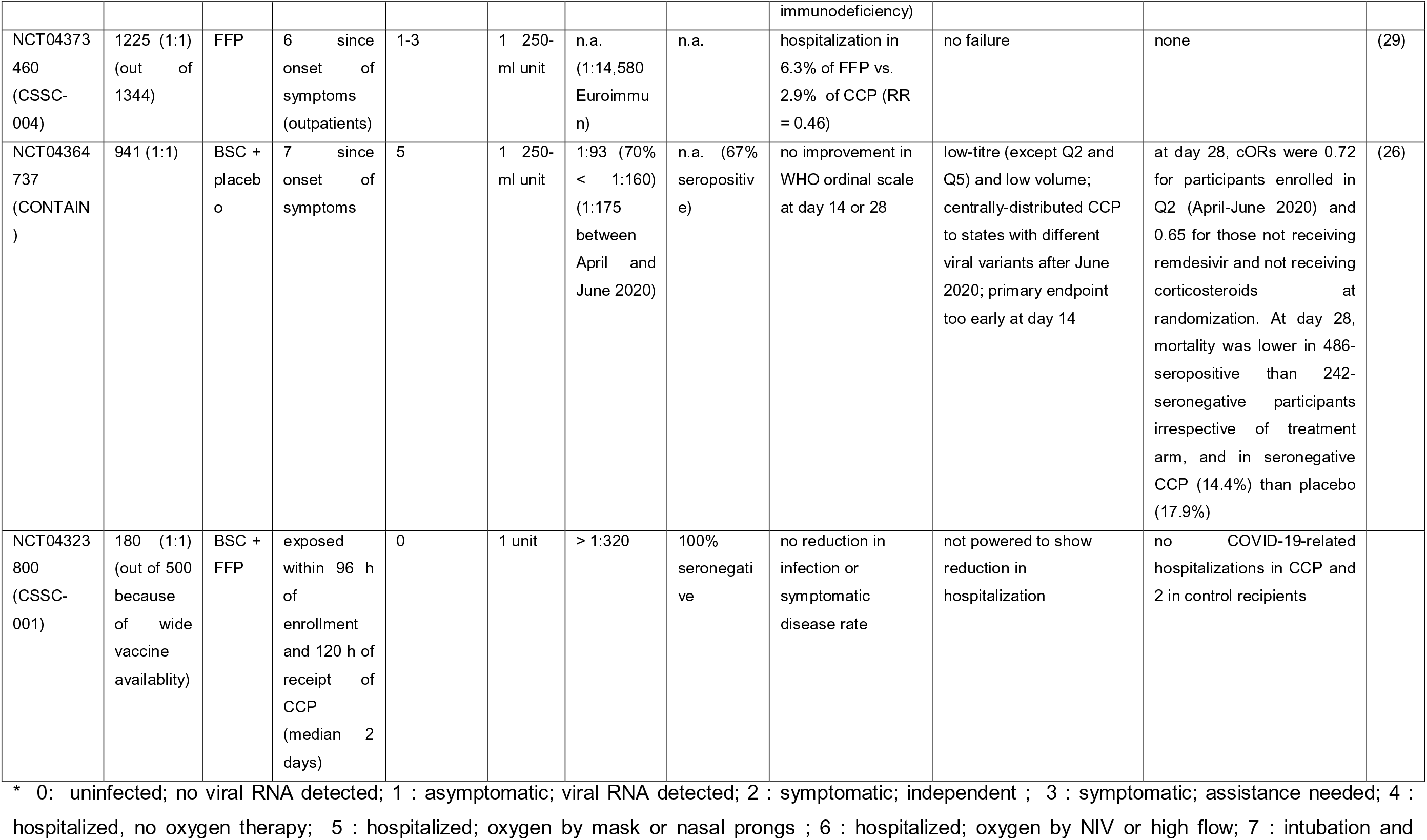
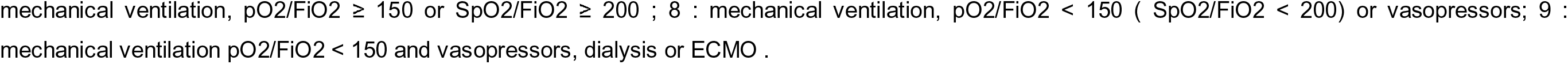
Randomized controlled trials (RCT) of COVID-19 convalescent plasma (CCP) reported to date, listed accoding to date of (pre-)publication. nAb: neutralizing antibodies. BSC: best supportive care. FFP: fresh frozen (nonconvalescent) plasma. n.a.: not assessed (i.e. antivirus antibodies were assessed only using high-throughput serology). IQR: interquartile range. Moderately late usage is defined as 4-6 days since onset of symptoms, late usage as 7-10 days, very late usage as >10 days.

**Table 3.**
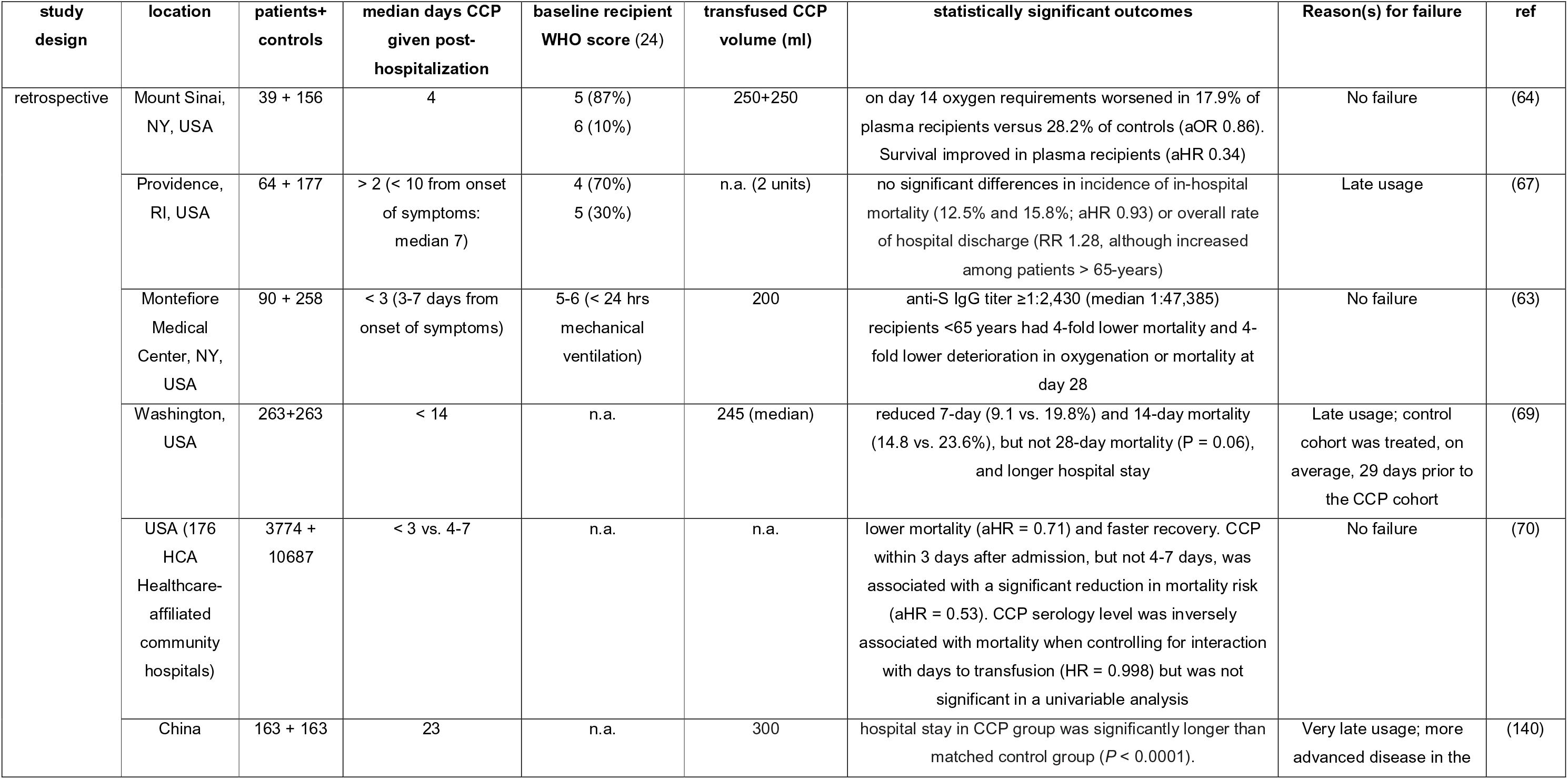
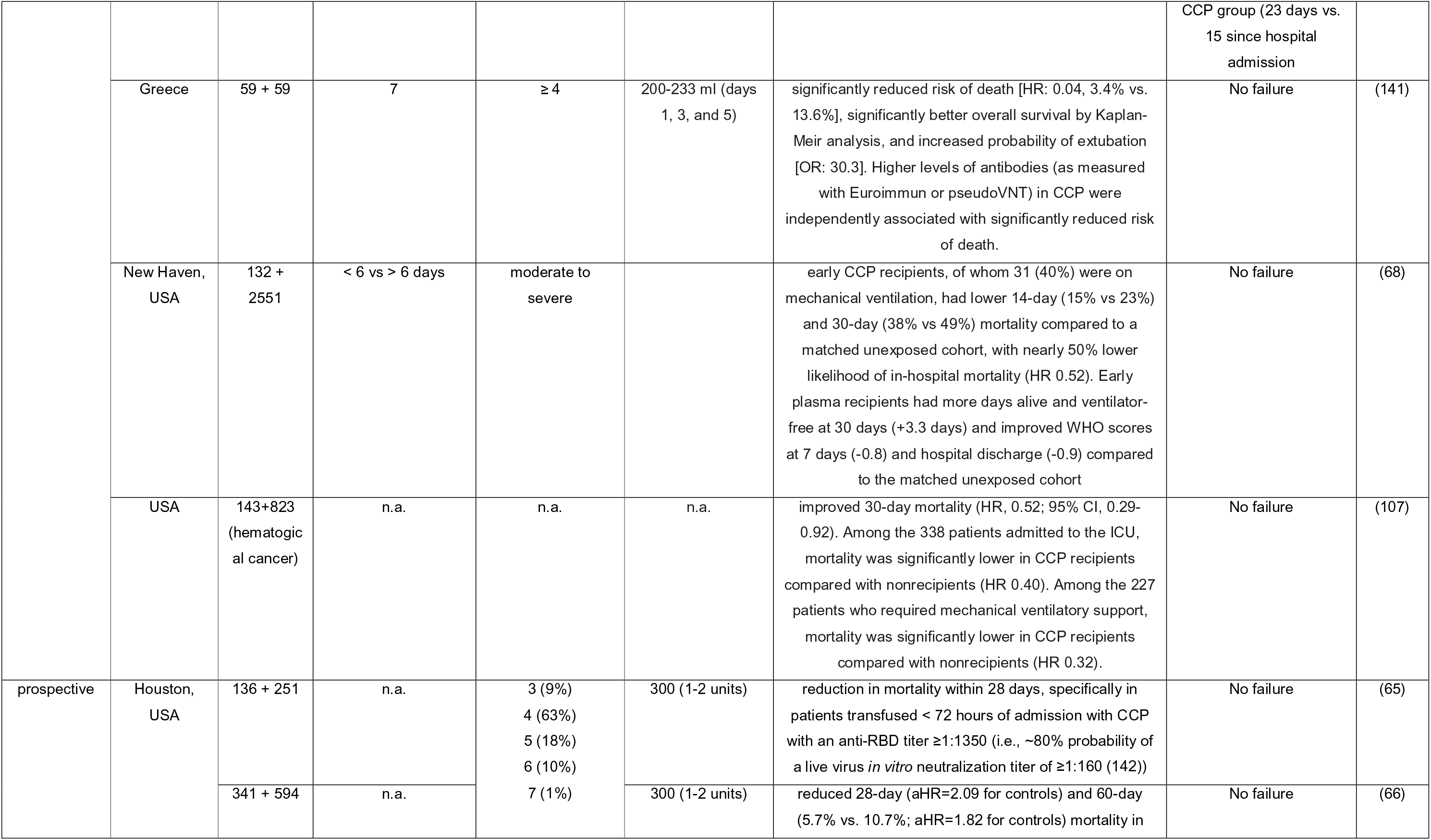
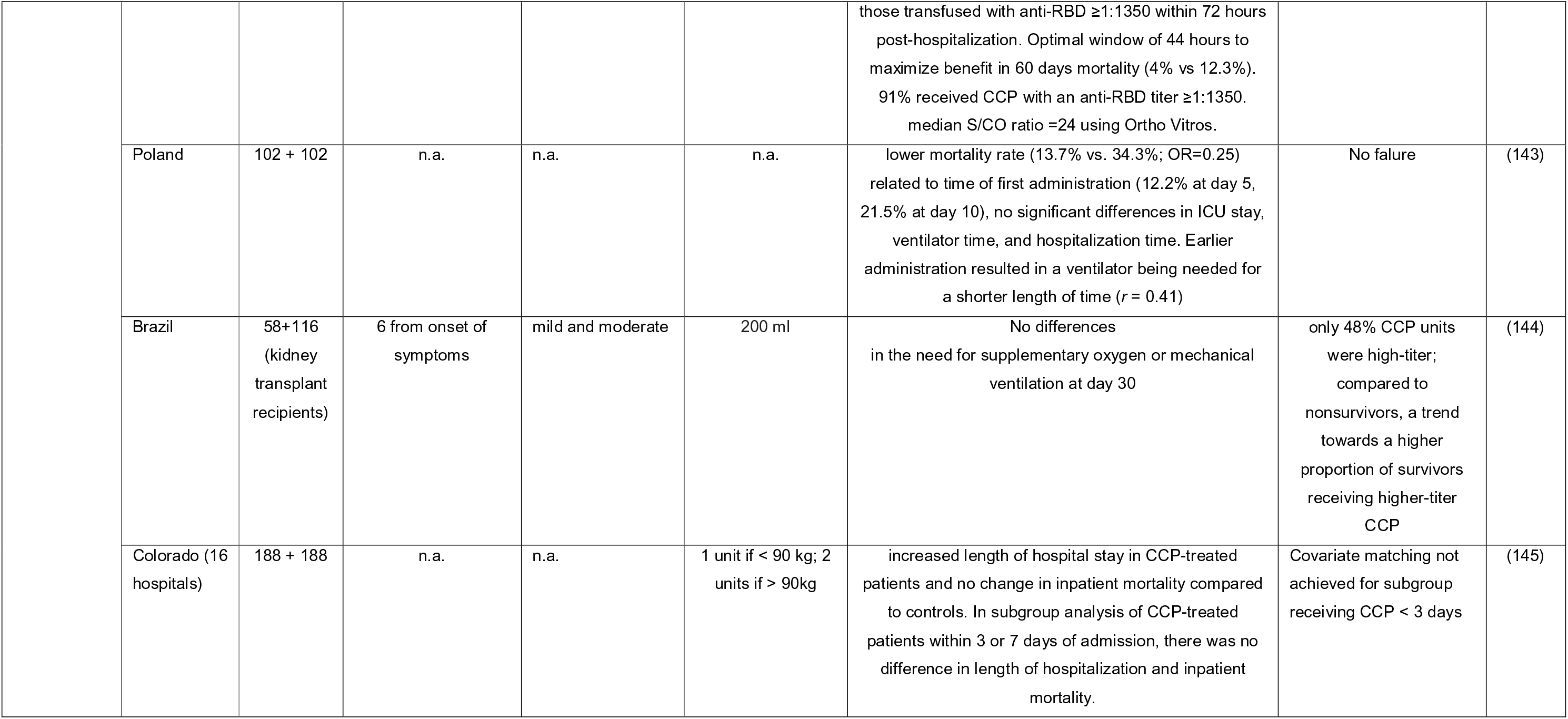
Propensity score-matched (PSM) CCP studies reported to date. DPH: days post-hospitalization. None of these studies titered nAbs in either donor or recipient using VNT. HR : hazard ratio ; OR : odds ratioL aOR : adjusted OR. aHR : adjusted HR.

ClinicalTrials.gov search retrieved 7 CCP RCTs completed but not yet prepublished or published, 5 active but not yet recruiting RCTs, and 12 RCTs which are still recruiting (summarized in Table 4).

**Table 4.** Summary of completed but not yet reported or ongoing RCTs of CCP, as registered on ClinicalTrials.gov as of August 26, 2021. BSC: best supportive care; FFP: fresh frozen plasma. Several studies were withdrawn (NCT04377568).

| Status | NCT number | Patient subtype | Control arm | Study design | Number planned to enrol | Study start | Locations |
| --- | --- | --- | --- | --- | --- | --- | --- |
| Completed | NCT04332835 | severe (SOFA <6) | BSC | single masking (outcomes assessor) | 92 | August 8, 2020 | Colombia |
|  | NCT04349410 | any | 10 different arms | single masking (investigator) | 1800 | April 11, 2020 | USA |
|  | NCT04421404 (CAPRI) | within 3 days from hospitalization or 14 from symptoms | placebo | triple masking (participant, care provider, investigator) | 34 | June 9, 2020 | USA |
|  | NCT04374526 | pneumonia, age > 65 and PaO<sub>2</sub>/FIO<sub>2</sub> ≥300 mmHg and comorbidities | BSC | open label | 29 | May 27, 2020 | Italy |
|  | NCT04358783 | hospitalized requiring supplemental oxygen | BSC | quadruple masking (participant, care provider, investigator, outcomes assessor) | 30 | April 27, 2020 | Mexico |
|  | NCT04405310 | moderate to severe requiring supplemental oxygen | albumin | double (participant, care provider) | 80 | May 20, 2020 | Mexico |
|  | NCT04425915 | on ventilator within 3 days from onset of symptoms | BSC | open label | 400 | June 14, 2020 | India |
| Active, not recruiting | NCT04539275 | ventilated and within 3 days from hospitalization | masked saline placebo | triple masking (participant, care provider, investigator) | 702 | November 16, 2020 | USA |
|  | NCT04374487 | hospitalized and severe | BSC | open label | 100 | May 9, 2020 | India |
|  | NCT04425837 | high-risk | BSC | single masking (outcomes assessor) | 236 | July 2020 | Colombia |
|  | NCT04395170 | hospitalized | 2 arms (BSC; anti-COVID-19 IVIG) | open label | 75 | September 2020 | Colombia |
|  | NCT04391101 | severe | BSC | open label | 231 | June 2020 | Colombia |
| Recruiting | NCT04516811 | moderate to severe | placebo | triple masking (participant, care provider, investigator) | 600 | September 21, 2020 | South Africa |
|  | NCT04388410 | hospitalized, severe disease or risk for severe diseases | BSC | quadruple masking (participant, care provider, investigator, outcomes assessor) | 410 | August 25, 2020 | Mexico |
|  | NCT04385043 | severe infection | BSC | open label | 400 | May 1, 2020 | Italy |
|  | NCT04380935 | acute respiratory distress syndrome | BSC | open label | 60 | May 18, 2020 | Indonesia |
|  | NCT04362176 (PassitOn) | hospitalized adults | placebo | triple masking (participant, care provider, outcomes assessor) | 1000 | April 24, 2020 | USA |
|  | NCT04390503 | exposed within 7 days or mild symptoms within 5 days | albumin | double masking (participant, outcome assessor) | 150 | March 12, 2021 | USA |
|  | NCT04376034 | severe or life-threatening | BSC | open label | 240 | March 30, 2021 | USA |
|  | NCT04366245 | hospitalized ventilated | BSC | open label | 72 | April 23, 2020 | Spain |
|  | NCT04333251 | hospitalized within 7 days from symptoms | BSC | open label | 115 | April 1, 2020 | USA |
|  | NCT04345991 | mild and within 8 days from symptoms | BSC | open label | 120 | April 15, 2020 | France |
|  | NCT04372979 | hospitalized within 10 days from symptoms | FFP | triple masking (participant, care provider, outcomes assessor) | 80 | September 14, 2020 | France |
|  | NCT04345289 | adults with pneumonia | placebo | quadruple masking (participant, care provider, investigator, outcomes assessor) | 1100 | May 1, 2020 | Denmark |

### The indication

While it would be desirable to have a single drug that works at any disease stage, it was not reasonable to expect neutralizing antibody (nAb)-based treatments such as CCP to have a significant effect in later stages of disease. COVID-19 is now well-defined as a disease with two stages, an initial viral phase characterized by flu-like and upper and lower respiratory symptoms, followed, in severe cases, by an inflammatory phase that is characterized by inflammation-driven damage to multiple organ systems, including the lungs that can impair gas exchange and cause life-threatening hypoxia and damage to multiple organs, including the brain and blood vessels (13). Accordingly, lack of endogenous nAb at baseline has been associated with a higher risk of viremia, but Marconato *et al* showed that CCP recipients benefit from high-titer CCP even after adjusting for their endogenous nAb (14). Specific intact antibodies in CCP are expected to neutralize SARS-CoV-2 in the intravascular system and, in some patients, prevent progression from early to severe and life-threatening disease (as seen in animal models (15–17)). However, this antiviral therapy cannot be expected to reverse the inflammatory phase of the disease, nor neutralize infectious viruses invading the extravascular system. Thus, COVID-19 is similar to influenza, a disease in which antivirals are effective early in disease but have no effect in later stages when the symptomatology stems largely from the inflammatory response. The rationale for administering CCP as early as possible in the course of COVID-19 stems from the neutralization stoichiometry itself: the larger the number of actively replicating virions in the body, the higher the nAb dose needed for neutralization (18). Some uncontrolled studies have reported a lack of association between early intervention and outcomes (19, 20), but in these studies the level of nAb or the overall anti-Spike antibody level in the infused CCP was unknown, leaving room for alternative explanations.

At the beginning of the pandemic, some investigators and opinion leaders, riding the wave of CCP successes in anecdotal reports in the media and small case series, introduced CCP to the general public as a panacea for any patient with COVID-19, including life-threatening cases, leading to confusing messaging: after reports of failure in severely ill patients emerged, opinions became polarized and the debate became anything but scientific (21). In clinical trials, the indication (i.e., the baseline clinical setting) has been variously defined by patient status (outpatient vs. presenting to the emergency room vs. hospitalized vs. ICU-admitted), disease severity (using 5-category COVID-19 Outpatient Ordinal Outcome Scale (22), a 6-category ordinal scale (12), a 7-category COVID-19 severity scale (23), the WHO 8- (24) or 11-category (25) ordinal scales, or pneumological scores such as SOFA), the time elapsed before recruitment (also variably defined as from molecular diagnosis, from onset of hospitalization, from diagnosis of pneumonia, or from onset of symptoms), or by serological status (presence of antibodies or the ability to neutralize SARS-CoV-2). This variability in inclusion criteria for studies has resulted in marked heterogeneity in recruited patients. As shown by CONTAIN (where those with shorter of symptom durations did worse), symptom duration can a poor indicator of ‘early’ disease but actually an indication of severe, rapidly progressive disease (26). Disease severity marked by WHO score as opposed to symptom duration may be a more accurate tool to capture “early” disease, as supported by the positive results with CCP in low WHO scores summarized in Figure 1.

**Figure 1.**
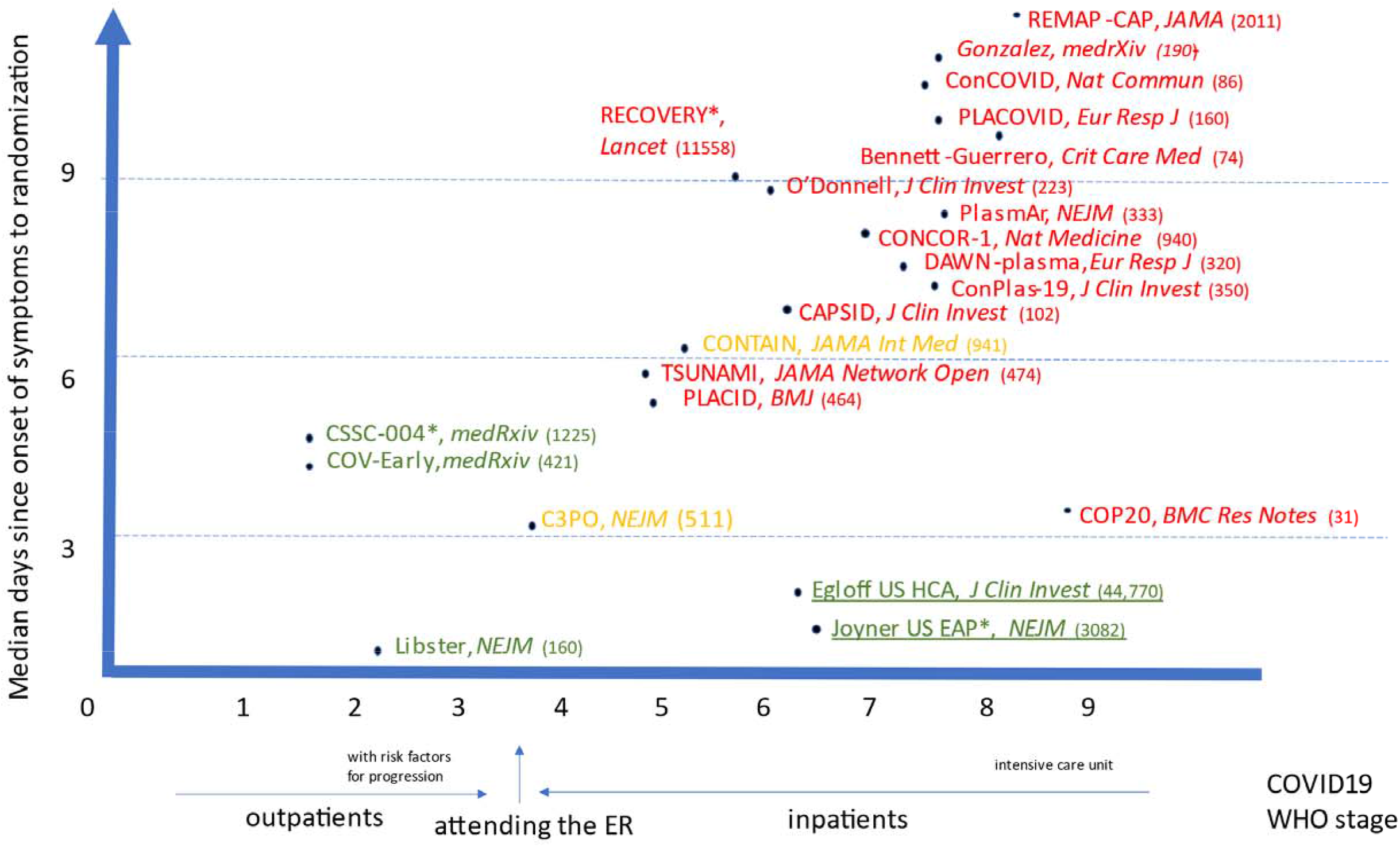
Simplified graphical representation of CCP RCTs reported ot date, plotted according to earliness of intervention and disease severity (stratified according to WHO 11-category ordinal scale (25): 0: uninfected; no viral RNA detected; 1 : asymptomatic; viral RNA detected; 2 : symptomatic; independent ; 3 : symptomatic; assistance needed; 4 : hospitalized, no oxygen therapy; 5 : hospitalized; oxygen by mask or nasal prongs ; 6 : hospitalized; oxygen by NIV or high flow; 7 : intubation and mechanical ventilation, pO2/FiO2 ≥ 150 or SpO2/FiO2 ≥ 200 ; 8 : mechanical ventilation, pO2/FiO2 < 150 (SpO2/FiO2 < 200) or vasopressors; 9 : mechanical ventilation pO2/FiO2 < 150 and vasopressors, dialysis or ECMO). Green text indicates trials which met the primary endpoint with statistical significance; orange text indicates trials which failed to meet the primary endpoint but showed statistically nonsignificant trends in favor of CCP; red text indicates trials which failed to show and benefit from CCP in the primary endpoint.

The vast majority of SARS-CoV-2 infections are mildly symptomatic, so when dealing with outpatients (WHO categories 1 to 3; the strata which are most likely to benefit from nAb-based therapies), the number needed to treat (NNTT) in order to prevent a single hospitalization or death can be very large, and even larger if vaccinees are recruited. In order to be economically and logistically sustainable, it seems wise to focus on those outpatients having risk factors for disease progression, as done with COVID19 mAbs. This approach was pursued by C3PO (27) and NCT04479163, but not other outpatient RCTs (COV-Early (28), CSSC-004 (29)). A special category is represented by outpatients recruited at the time of ER attendance (e.g. in C3PO (27): they should be considered at the border between outpatients and inpatients, and hence not aggregated in outpatient metanalyses. Among outpatients not recruited at the ER, clinical benefit has been shown up to 6 days since onset of symptoms (e.g. in the CSSC-004 RCT (29)). No benefit of CCP over FFP has been proven in the single RCT of post-exposure prophylaxis (CSSC-001) at preventing infection, symptomatic disease, but the study was not powered to detect a reduction in hospitalization (30).

For what concerns inpatients (WHO categories 4 to 9), clinical benefit from antivirals can only be expected for categories 4 and 5, since from category 6 on the patients require high-flow oxygen and hence have significant respiratory failure.

### The therapeutic dose

Determining the effective dose of CCP is difficult in a pandemic because the antibody assays and other tests needed to assess the potency of any antibody product take time to be developed. In practice, the effective dose is the product of multiple factors, none of which is fully standardized.

The first factor is the concentration of the nAbs as measured by a viral neutralization test (VNT). At the beginning of the pandemic, only a few biosafety level 3 (BSL3) (or higher)-equipped virology laboratories could run VNT using authentic live SARS-CoV-2 virus: the procedure was time-consuming (3-5 days) and the reports were operator-dependent. Nowadays, the availability of Spike-pseudotyped viruses which can be managed under the more widely available BSL-2 laboratories, or cell-free ACE-2 competition assays, combined with automated (e.g., luminescence-based) readings, have standardized outcomes and shortened turnaround times (31): however, harmonization between different assays is still a work in progress (32). The VNT differs according to the type of replication-competent cell line, the viral isolate used for the challenge (which is critically important when the virus is mutating rapidly as has been the case with emergence of variants of concern), the multiplicity of infection (i.e., the ratio between the viral inoculum - referred with different measuring units – and the number of replication-competent cells within each well), the detection system (optic microscopy for cytopathic effect, immunostaining, quantitative PCR, or luminometer for engineered pseudoviruses), and finally the threshold of neutralization (50% or 90%). The DAWN-plasma (33), C3PO (27), and REMAP-CAP (34) RCTs provide clear examples of such heterogeneity, with up to 4 different VNTs used at different participating laboratories/countries within the same study (see Table 1). It was not until August 2020, when many trials were already underway, that the FDA Emergency Use Authorization 26382 defined high-titer CCP on the basis of correlation with a reference standard, the Broad Institute live-virus, 5-dilution VNT as a 50% inhibitory dilution (ID_50_) of 1:250 or more (https://www.fda.gov/media/141481/download), and exclusive use of high-titer CCP was formally recommended by the FDA only on March 9, 2021.Table 1 summarizes the key variables in VNT employed to date in CCP RCTs. Published trials have varied greatly in their approaches to antibody quantification whether in measured transfused CCP units or in recipients. Several RCTs performed nAb titration, but with highly heterogenous methods which makes comparability of doses across studies difficult. Table 1 attempts to reconcile doses across those trials, showing that they actually differed more than is apparent by inspection of raw titers.

Despite these uncertainties, we can make estimates of likely effective doses based on the available clinical experience thus far. The lack of utility from low-titer (1:40) CCP in moderate COVID-19 was confirmed by the PLACID trial (10). As long as a clear therapeutic dose is not identified, it seems prudent to transfuse units containing nAb titers at least 10-fold higher than the nAb titer measured before transfusion in recipient serum. Similarly, the ConCOVID RCT showed that CCP units having nAb titers similar to those of the recipients (1:160) did not confer a clinical benefit (35). CCP units with an adequate nAb titer (nowadays estimated at ≥1:160) are more easily found among older males who recovered from a previous symptomatic COVID-19 requiring hospitalization (36, 37): unfortunately, such donors were poorly represented in the first donation waves, which tended to obtain CCP from younger donors will mild disease, and, presumably lower nAb titers (10).

Many trials have relied on high-throughput semi-quantitative or qualitative assays with a poor-to-moderate relationship with nAb titers. Harmonization of such high-throughput assays using the WHO International Standard of binding arbitray units (BAU) is nowadays possible (38). Although most trials performed a correlation analysis between VNT and high-throughput serological assays, in many cases the CCP units were tested only with the latter without validation, as was the case with 66% of the patients in the PlasmAR trial (12). This procedure risks an incorrect evaluation of the neutralizing CCP activity. Another cause for discrepancies in outcomes could be that although IgM, IgG, and IgA are all capable of mediating neutralization, VNT titers correlate better with binding levels of IgM and IgA_1_ than they do with IgG (39). By contrast, routinely used high-throughput serological assays only quantify IgG, including non-neutralizing IgGs, whose potency against SARS-CoV-2 has not been established. Trials should preferentially use VNTs to assess serostatus of transfused units and not rely on high-throughput serology. As for any other medicinal product, CCP exhibits a dose-response relationship, which is also evident when using high-throughput assays. In the subgroup analysis of the EAP, a gradient of mortality was seen in relation to IgG antibody levels in the transfused CCP. In the subgroup of patients who were not receiving mechanical ventilation, death within 30 days after CCP transfusion occurred in 81 of 365 patients (22.2%; 95% CI, 18.2 to 26.7) in the low titer group, 251 of 1297 patients (19.4%; 95% CI, 17.3 to 21.6) in the medium-titer group, and 50 of 352 patients (14.2%; 95% CI, 10.9 to 18.2) in the high-titer group. Depending on the statistical model, the RR for 30-day mortality in high-titer CCP compared to low-titer CCP recipients ranged from 0.64 – 0.67, with an upper 95% confidence bound of 0.91 (8). Similarly, the large retrospective PSM study from Hospital Corporation of America (HCA) Healthcare-affiliated hospitals reported a 0.2% decreased risk of mortality for every 1 unit of S/Co serology level (40).

An additional limit is testing nAbs only at the time of first donation (e.g., in ConCOVID (35)) while leaving donors opportunities to repeat donation for months: given the expected decline of nAb levels in time in convalescents, this could have overestimated the actual dose.

The nAb titer (or total IgG levels as measured by surrogate assays) only describes one factor involved in defining the real therapeutic dose in that it represents the concentration of just one (likely the main) active ingredient. But CCP contains additional antibodies that mediate antibody-dependent cellular cytotoxicity (ADCC), complement activation and phagocytosis of viral particles, functions that can each contribute to its antiviral effects (41). At this time the relative importance of nAbs vs. the other antibody activities is not understood, but, hopefully, retrospective analyses that correlate CCP efficacy with these activities will reveal additional variables that need to be considered in choosing optimal CCP units. This has relevance when assessing interfering factors: e.g., the impact of pathogen reduction technologies (PRT) on nAb has experimentally been shown to be apparently minimal (42, 43), but their impact on Fc-dependent functions (such as ADCC) is still to be investigated and could be relevant for some PRTs (44) (used e.g. in TSUNAMI (45), NCT04356534 (46), and ConPlas-19 (47). In this regard, we note that an early study reported that methylene blue reduced the protective function of antibodies to pneumococcus by interfering with glycosylated domains, raising concerns that it could affect Fc function (44). Of relevance, the various pathogen-reduced plasma went through clinical evaluation to assess their hemostatic activity linked to coagulation factors, or, at best, Fc binding to receptors (48), not their immunological activity : furthermore, none of the photochemical processes used for CCP is used in the field of plasma fractionation, and therefore we cannot make inferences from pharmaceutical-grade immunoglobulins.

The therapeutic dose of nAb is a product of its concentration in the infused CP multiplied by the overall infused CP volume, adjusted to the recipient body weight to take account of dilution into the blood volume and tissues. RCTs have varied in the provision of volume per unit (200-300 ml), and most importantly in cumulative volume per patient (1-4 units) and in extent of exposure to diverse antibodies from various CCP donors, and no published trials have adjusted levels of nAbs by recipient body weight (or, when attempts have been performed, they referred to the historical 10-15 ml/kg dose derived frm the treatment of hemorrhagic coagulopathies (49)). A failure of CCP to improve outcomes when 200-ml of 1:160 nAb-titer CCP is provided to a patient who weighs 120 kg represents quite a different scenario from failure of a 600-ml transfusion of 1:640 nAb-titer CCP to produce improvement in a 60-kg patient. But these central issues in dosage have not been considered in the RCTs published so far.

Finally, antibodies other than nAbs can play a prominent role in viral clearance. Bahnan *et al* have shown that CCP and anti-Spike mAbs induce phagocytosis but with diminishing returns when the antibody concentrations become high: activation and inhibition of phagocytosis are independent of neutralization potential and humanized ACE2 mice are protected from intranasal challenge by non-neutralizing antibodies (50).

### Relevance of CCP to the viral variant

Albeit not formally demonstrated, CCP manufactured by pooling ABO-matched units from many different donors (e.g., in PlasmAr (12)) theoretically have greater polyclonality of nAbs than repeated CCP doses from a single-donor (e.g. CAPSID (51)) and should grant higher efficacy against viral variants. Nevertheless, pooling typically occurs among donors attending the same blood bank, making donor exposure to different viral variants unlikely.

An analysis of potential variables associated with CCP efficacy associated near-sourcing with reduced mortality, with the efficacy of CCP in reducing mortality falling sharply when the CCP source was more than 150 miles from where it was used (52). This finding suggests that SARS-CoV-2 variants atsome geographic locations create antibody responses in CCP that are not effective against other variants at different locations (53, 54). Even though CCP is often standardized for nAb titer to the Spike protein, the VNT could use a nonrelevant viral strain, or miss major functional differences for the antibody response (41). This finding has implication for RCTs that use nationally sourced (centralized) CCP, since the attempt to standardize the therapeutic units centrally could inadvertently reduce CCP efficacy if hospitals use CCP obtained from distant loci. For example, in the C3PO RCT, which was conducted in 21 USA states, 95% of the donor CCP was collected in either Chicago or Denver: since only 4 of the 48 centers were in Illinois or Colorado, most CCP usage had to be from remote sources (27). The same concern applies to large multinational RCTs such as REMAP-CAP (34). By contrast, the NCT04359810 RCT in New York and Brazil, which found a beneficial effect of CCP on mortality, used CCP locally sourced in New York, whose efficacy against P.1 was tested to ensure efficacy at the other recruiting center in Brazil (11).

Although also not formally demonstrated during clinical trials, it is also reasonable to assume that CCP collected during early pandemic waves could be less effective against currently circulating variants of concern (55). RCTs whose recruitment was protracted across multiple pandemic waves (e.g., ConPlas-19 (47) and TSUNAMI (45)) and which relied on CCP collected and banked months earlier could have inadvertently used CCP with reduced activity against the SARS-CoV-2 strains circulating the community when the therapy was administered. No CCP study to date has ever annotated or retrospectively performed SARS-CoV-2 sequencing on CCP donors nor on patients to identify the underlying variant: matching of the convalescent donor and the recipient for viral variant has neber been formally achieved. Hence, both geography and time of collection of the CCP are important variables when considering the efficacy of the treatment.

### The intended outcomes

Most trials (CONTAIN (26) and PassItOn being exceptions) have used composite endpoints or specialty scores (e.g., SOFA) rather than progression in the simple WHO ordinal scale or mortality, and many were stopped because of apparent futility at a time when they may have been underpowered to detect significant benefit. As represented in Figure 2, several studies have reported overall negative results (panel A) despite the presence of positive signals of efficacy just barely missing statistical significance (panels B and C). The significance level (i.e., *p*= 0.05) is largely a socially constructed convention for rejecting the null hypotheses, but it has often been misinterpreted as a measure of reality by many individuals not familiar with the nuances of statistics. For example, some CCP studies have concluded that a difference that did not achieve a p value < 0.05 was an absence of difference, even when mortality in the CCP arm was ∼20-40% lower than in controls (11, 47, 51, 56). This reasoning has played a central role in the polarized views of CCP efficacy and has impeded subsequent studies from drilling down on positive effects that were observed. The dogged pursuit of statistical significance, viewed as a measure of reality instead of the actual reality demonstrated by the data, during a public health emergency dealt a serious blow to studies of CCP and created significant confusion for clinicians. It is also important to understand that RCTs are powered to be less tolerant of Type I error than Type II error, which are conventionally set at .05 and .20, meaning that a Type II error is expected four times as often as a Type 1 error. This statistical convention can contribute to the absence of significance in studies that were set up early in the pandemic when there was little information on expected effects for the various patient populations studied, when power estimates were only guesses, and the enrolled patients were so heterogenous that only subgroups were likely to have responded to CCP treatment. Many studies were originally designed to enroll inpatients at any disease stage, and it should be no surprise that subgroup analyses on the groups that were later demonstrated to be more likely to benefit from CCP (e.g., early treated, seronegative patients, those receiving high nAb titre) were underpowered to reach statistical significance, as shown by orange color predominance in panel C of Figure 2. Nevertheless, favorable trends are a shared feature across such trials (5, 11, 12, 27, 47, 49, 51, 56, 57).

**Figure 2.**
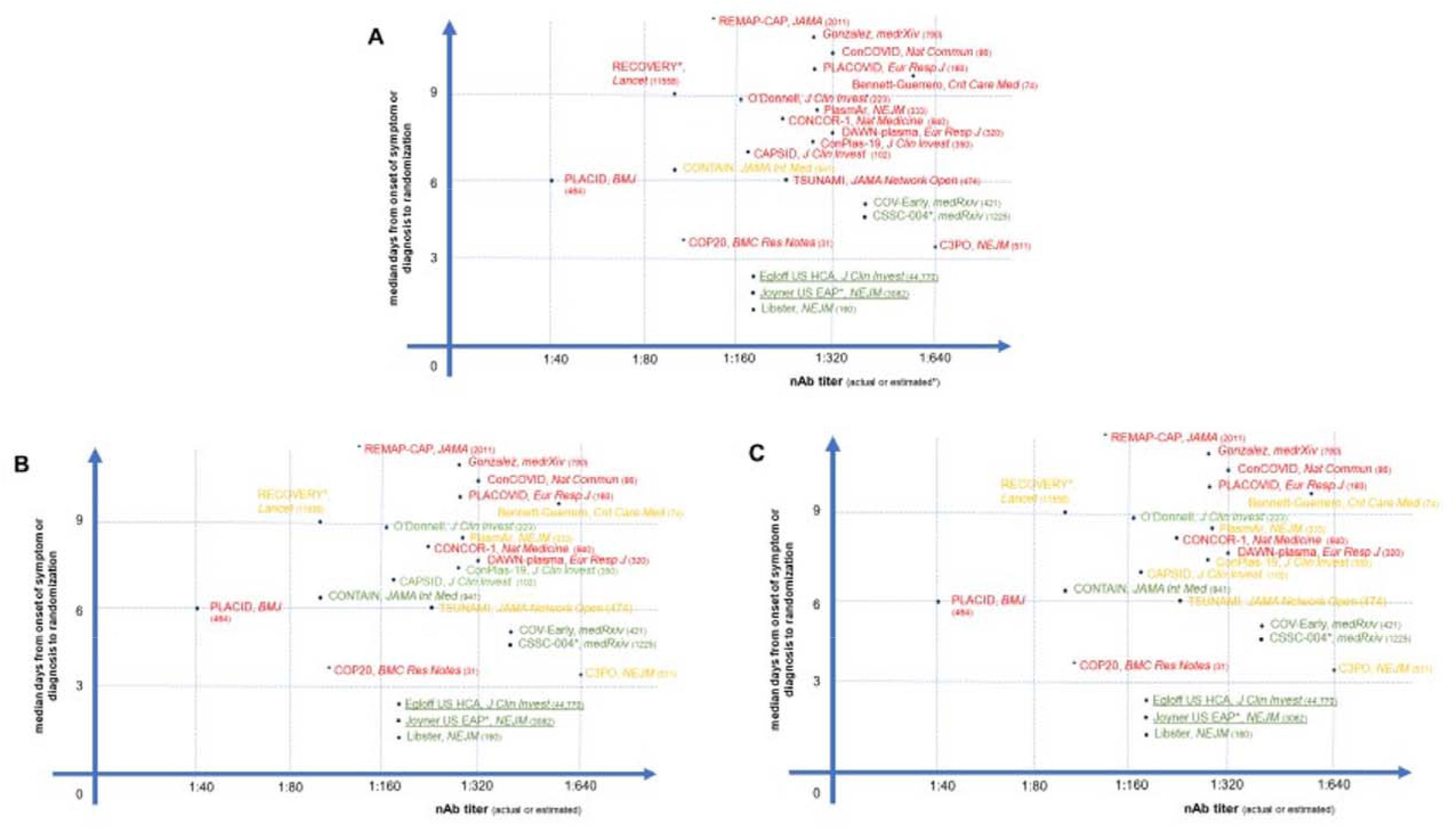
Simplified graphical representation of CCP RCTs reported to date, plotted according to earliness of intervention and nAb titers in CCP. In **panel A**, green text indicates trials which met the primary endpoint with statistical significance; orange text indicates trials which failed to meet the primary endpoint but showed statistically nonsignificant trends in favor of CCP; red text indicates trials which failed to show and benefit from CCP in the primary endpoint. In **panel B**, green text indicates trials which showed overall mortality benefit from CCP; orange text indicates trials which showed mortality benefit from CCP in the subgroup of early arrivals or higher nAb titers; red text indicates trials which failed to show any mortality benefit from CCP. In **panel C**, green text indicates trials which showed statistically significant mortality benefit from CCP (overall or in the subgroup of early arrivals or higher nAb titers); orange text indicates trials which showed statistical trends towards mortality benefit from CCP (overall or in the subgroup of early arrivals); red text indicates trials which failed to show any mortality benefit trend from CCP in any subgroup. Underlined text indicates large trials which were not RCT and for which nAb levels was inferred from high-throughput serology, but are nevertheless reported as reference studies. Numbers in parenthesis indicate cumulative number of patients enrolled.

Rrigid adherence to primary outcomes that were often fixed in the early days of the pandemic when information about disease stage and quality of CCP associated with efficacy were not understood has contributed further to the confusion. When these outcomes were not met, trials were considered failures even though there were often signals of efficacy in the data that were not considered as valuable since these had not been pre-specified, even when they made biological sense. For example, in the New York-Brazil RCT cited above, CCP did not lower the primary end-point of clinical status on an ordinal scale, but the statistically significant halving of mortality was acknowledged in the abstract. Would it have made sense to ignore the strong effect of CCP on mortality in this trial just because mortality was not selected as a primary outcome? RCTs of mAbs (58) and dexamethasone (59) were designed to achieve a tiny 6% and 4% reduction in mortality, respectively, while several CCP RCTs failed because their initial assumption in the magnitude of reduced mortality was more optimistic (e.g. TSUNAMI (45)).

Another misunderstood endpoint is viral clearance, defined as the conversion of nasopharyngeal swabs (NPS) from positive to negative for PCR evidence of SARS-CoV-2 in CCP-treated patients. While there was early and robust evidence for this effect from CCP (4, 10), sampling NPS too late after CCP treatment, when the endogenous immune response had also mounted in the control arm, could miss differences.

### Analyzing conflicting outcomes in individual RCTs

We use the word ‘failures’ with care and considerable nuance, since negative trials can be very important in teaching us about populations that do not benefit from CCP or variables that affect its efficacy. Keeping the factors discussed above in mind, we have analyzed individual RCTs in detail. At the very beginning, many historically or internally controlled observational studies showed clinical benefit from CCP (4–6) and this led the FDA to issue an EAP in March 2020 that was converted into an emergency use authorization (EUA) in August 23, 2020. The largest observational study is the US open-label EAP (NCT04338360) led by Joyner *et al*, which enrolled 105,717 hospitalized patients with severe or life-threatening COVID-19 from April 3 to August 23, 2020 (60). In an analysis of the effect of antibody in CCP performed independently of the results cited above (8) and using a nAb titer in an overlapping but non-identical group of EAP patients, the FDA showed that the 7-day mortality in non-intubated patients who were younger than 80 years of age and were treated within 72 hours after diagnosis was 6.3% in those receiving high-titer CCP and 11.3% in those receiving low-titer CCP (https://www.fda.gov/media/142386/download).

In a later analysis of a larger (N = 35,322) subset of EAP patients, (including 52.3% in the intensive care unit (ICU) and 27.5% receiving mechanical ventilation), the 7-day mortality rate was 8.7% in patients transfused within 3 days of diagnosis but 11.9% in patients transfused ≥4 days after diagnosis; similar findings were observed in 30-day mortality (21.6% vs. 26.7%) (61). In an EAP study from Argentina, mortality after CCP was 18.1% among 3,113 patients treated within 3 days since hospitalization, 30.4% among 1380 patients transfused between days 3-7, and 38.9% among 226 patients transfused after day 7 (62). The major criticism of these results is that controls were neither randomized nor PSM: hence a difference in the treatment outcome between treated and untreated groups may be caused by a factor that predicts treatment rather than by the treatment itself. However, importantly, nAb titer analysis was retrospectively done, both patients and physicians were unaware of the nAb content in the CCP units used, the results are what would have been expected from the experience with antibody therapy, and multivariate models were used to adjust for potential confounders (1). Additionally, given the outline of an optimal use case with this data and the earlier underpowered RCT by Li *et al* (5), it is unfortunate that due to (a) lack of awareness and (b) logistical burden associated with protocol adjustments, involving repowering and new patients’ recruitment criteria, later treatment RCTs either continued or initiated without modifications to include newly available evidence.

The highest level of scientific evidence in primary clinical research stems from prospective PSM and RCTs. PSM studies (Table 3) balance treatment and control groups on a large number of covariates without losing a large number of observations. Unfortunately, no PSM study to date has investigated nAb titers by VNT or outpatients. Nevertheless, in 2 retrospective PSM studies from 2 different hospitals in New York, trends for improved outcomes in non-intubated and those treated within 7 days since hospitalization (HR 0.33) were observed (63, 64). These findings were later confirmed in a prospective PSM study from Houston (65, 66). Of interest, a retrospective PSM study from Providence did not show any benefit, but patients were treated at a median of 7 days after onset of symptoms (67). Another PSM study from Yale associated CCP with a 35% reduction in mortality (68). That study is notable in that it included patients on mechanical ventilation who would not normally be expected to benefit from CCP and the percentage of individuals receiving corticosteroids was very low since the study was conducted in the early days of the pandemic in the USA. Another PSM from the Washington DC area found a reduction in mortality with CCP use at both days 14 and 28, which reached statistical significance at the earlier date (69). Finally, a very large study from 176 community hospitals affiliated with HCA Healthcare confirmed substantial mortality reduction in hospitalized patients receiving CCP within 3 days from admission (70).

Since PSM only accounts for observed (and observable) covariates, and not latent characteristics, RCT remains the gold standard for highest-level evidence (Table 2). In the PlasmAr RCT, the small number of early arrivals (less than 72 hours) showed superior primary and secondary outcomes in the CCP arm (n= 28) compared to the placebo arm (n=11), but the minimal contribution of this group to the overall cohort (228 CCP and 105 placebo) made the advantage disappear in the final outcomes at day 30 (12). In another Argentinean RCT on 160 outpatients older than 65 years of age with mild COVID-19 who were treated with CCP within 72 hours, progression to severe COVID-19 halved at day 30 (71). Similar findings were reported in outpatients without risk factors in CSSC-004 (29). An RCT from India reported that patients younger than 67 treated at a median of 4 days after hospital admission manifested superior mitigation of hypoxia and survival in the CCP arm (72). Another RCT in Spain enrolling patients at less than 7 days of hospitalization showed four deaths in the control arm, none in the CCP arm (47). Similarly, the ConCOVID RCT showed reduced mortality in the CCP arm (35).

An additional complexity in recruitment to CCP trials is time to treatment. Clinical trials involve administrative requirements and consent procedures, and recruitment to a RCT further requires randomization, which may produce delays in treatment. CCP therapy requires cross-matching of blood types, ordering the CCP, which may or may not be available on site, and setting up the transfusion. This inherent delay from randomization to infusion means that RCTs may build in a disadvantage for the CCP study arm in the few RCTs using control treatments (e.g. saline or fresh frozen plasma (FFP)), where controls may have received treatment earlier in the disease course : for example, FFP was used in NCT04346446 (57), NCT04359810 (11), NCT04344535 (56), COV-Early (28), and CSSC-004 (29) RCTs. ABO-compatible CCP units may be not readily available at the local blood bank and recruited patients may have to wait for a compatible unit of CCP. These almost inevitable delays from randomization mean that CCP may be provided later in the illness than is ideal, and even if the trial intends to treat early, in practice it may not be possible unless the RCT is designed to deliver plasma immediately after randomization.

During a pandemic, moreover, delays in treatment are magnified. The accrual of severely ill patients in emergency departments and the overwhelmed or even collapsed health care systems can create long delays from arrival in the emergency room to treatment. In the absence of quick (antigenic or molecular) tests for SARS-CoV-2, the turnaround time for final confirmation of diagnosis with PCR, which must often be run in batches, can take several hours. All these factors are likely to impact the efficacy of CCP treatment. To shorten such time, fully screened CCP collected from eligible donors (73) could be safely administered within emergency departments shortly after admission and even before the patient reaches the ward.

Figure 2 graphically places the outcomes of RCTs and PSM studies on a Cartesian plot having timeliness and nAbs dose as variables (if values are disclosed in the reports): this makes immediately clear that the few successes at reaching the primary endpoints have gathered into the lower right corner (high nAb dose and early intervention), while the many conflicting outcomes have been scattered all around (panel A), reflecting lower antibody levels infused or late treatment, or both, with the latter being the commoner problem. Nevertheless, when we focus on mortality irrespective of statistical significance (panel B) or focusing on statistical significance (panel C), many more RCTs showed clear benefits.

We will focus here on “failures” as identified by title, abstract and/or press recognition. Narratively, we could group so-called “failures”, with failure implying inability to demonstrate a favorable outcome to CCP use, into 4 categories, according to the main reasons:

1. Trials that transfused insufficient therapeutic doses of CCP due to either low total IgG levels or low nAb levels (e.g., PLACID (10))
2. Trials that transfused appropriate doses of CCP but too late, but which nevertheless reported signals of efficacy (e.g., RECOVERY (49), CAPSID (51), NCT04359810 (11) and TSUNAMI (45))
3. Trials that were stopped too early to observe benefit or with inherent design flaws, and/or were underpowered such that likelihood of success was reduced (e.g., C3PO (27))
4. Trials in which CCP was used to treat a condition not amenable to antibody intervention, such as hypoxia that is caused by pulmonary inflammation (e.g., COP20 (74) or REMAP-CAP (34))

### The inadequacy of meta-analyses

With all the heterogeneity in key drivers discussed in the former paragraphs, it becomes clear that secondary research (ranging from umbrella reviews to meta-analyses to systematic reviews), whereby each study is considered at the same level, invariably ends up with biased and divergent conclusions. This adds confusion to the already complex field of individual trial outcomes. Amazingly, as of December 21, 2021, PubMed has indexed 26 meta-analyses on CCP efficacy, more than the RCTs reported at the same date. Until the beginning of 2021, meta-analyses (variably including observational studies) were generally in favor of CCP (75), but began to be biased towards failure after publication of the large RECOVERY trial (49), which, by enrolling as many as 11,448 patients, diluted all the signals from positive RCTs. Clear examples of this phenomenon come from a widely cited metanalysis from Janiaud *et al* in *JAMA* (76) which included press release data from RECOVERY, and from the living systematic review by the Cochrane Group (77). The *JAMA* paper was surely unprecedented in the tradition of meta-analysis, not only because it included a study based only on a news release (which proved to differ in some important respects from the published paper), but because it allowed these data from a news release to dominate the entire analysis. At November 2021, in a follow-up metanalysis from the same group on 16,477 patients from 33 trials, RECOVERY still weighted 69.8% (78). Several groups attempted to dissect the RECOVERY trial and others by running subgroup analyses in their systematic reviews (79–81), but these reviews were unable to restore confidence in CCP efficacy in the clinical community that had been lost because of the publication of the overall negative findings of RECOVERY and PlasmAr (82). Merely following such metanalyses, on December 7, 2021 the WHO revised its living guidelines on drugs for COVID19, discouraging usage of CCP (83): such contraindication received large criticisms from both the SUPPORT-E (84) and CCPP19 consortia.

Actually, metanalyses embedding more recent studies and performing subgroup analysis were more positive on the beneficial effects of CCP. A metanalysis of 22,591 patients (enrolled in 10 RCTs and 15 observational studies) showed that early CCP significantly reduced mortality (RR 0.72, p<0.00001), but only in patients who were not suffering severe or critical disease (85). Another metanalysis of 18 peer-review clinical trials, 3 preprints, and 26 observational studies actually found that CCP use was associated with reduced risk of all-cause mortality in severe or critical COVID-19 patients (86). A recent umbrella review of 29 metanalyses and systematic reviews found evidences for improvement in the CCP arms for some outcomes (overall mortality, viral clearance at day 3,) but not for others (clinical improvement, length of hospital stay (87).

Rather than pooling published RCTs, the Continuous Monitoring of Pooled International Trials of Convalescent Plasma for COVID-19 Hospitalized Patients (COMPILE) study pooled individual patient data from ongoing RCTs at two-week intervals. Unfortunately, with the single exception of CONTAIN (26), participating RCTs largely shared usage in advanced disease stages (DAWN-plasma (33), PLACID (10), ConCOVID (35), ConPlas-19 (47), NCT04421404, PennCCP2 (88), and the Brasília Covid-19 Convalescent Plasma (BCCP)) (89).

Current clinical utility of CCP While CCP contains a plethora of biologically active molecules (90), we now have very strong evidence that appropriately vetted CCP from eligible convalescent donors is safe for patients (91, 92), with no evidence of increased risks of transfusion-transmitted acute lung injury, antibody-mediated enhancement concerns feared in the early days of the pandemic (93) nor is there evidence that CCP induces accelerated SARS-CoV-2 evolution (11). Polyclonal antibodies such as CCP are likely to offer better protection against onset of variants than monoclonal antibodies (94–96): importantly, Pommeret *et al* showed that CCP can rescue immune escape variants emerged during treatments with bamlanivimab/etesevimab cocktail (97). Outcomes in immunocompromised patients treated with CCP have been successful in the long-term, with minimal evidence for immune escape (98).

We have also learned that CCP is less likely to benefit patients requiring oxygen (i.e., from level 4 and up on the 11-point WHO ordinal scale), and hence, ideally, the focus should be on outpatients and in identifying that subset of patients who seek hospital care and are still sufficiently early in the course of disease such that they can benefit from CCP. This finding parallels the finding with hyperimmune serum and anti-Spike monoclonal antibodies, which at first failed in hospitalized patients (99, 100), but later succeeded for ambulatory patients with mild to moderate COVID-19 (101) and were approved for emergency use. However, at this moment clinical use in the US is restricted by the FDA to inpatients.

CCP usage per admission peaked after issuance of the EUA, with more than 40% of inpatients estimated to have received CCP between late September and early November 2020. Oladunjoye *et al* showed that mortality in the second wave in USA, when utilization of corticosteroids, remdesivir and convalescent plasma was higher, was lower than the first wave (102). However, following reports of RCTs that failed to show clear benefit from CCP, usage per admissions declined steadily to a nadir of less than 10% in March 2021. A strong inverse correlation (Pearson correlation coefficient of -0.5176 with *P* = 0.00242) was found between CCP usage and deaths occurring 2 weeks after admission, and this finding was robust to examination of deaths taking place 1, 2 or 3 weeks after admission. Changes in the number of hospital admissions, prevalence of variants, and age of patients could not explain these findings. The authors estimated that the retreat from CCP usage, a phenomenon they termed “plasma hesitancy”, might have resulted in 29,000 to 36,000 excess deaths in the period from mid-November 2020 to February 2021 (103). The same analysis estimated that USA had avoided 96,000 excess deaths from August 2020 to March 2021 by its liberal deployment of CCP.

Several lines of evidence, ranging from the EAP to clinical trials employing RCT or PSM controls are now indicating how CCP should be used in immunocompetent patients (104). The evidence supports the initiation of CCP treatment as early as 44-72 hours within onset of symptoms (which largely pertains to outpatients) and using CCP with a nAb titer > 1:160. Benefit within 1 week from onset of symptoms (including in hospitalized patients) is less well understood, although a benefit from higher therapeutic doses cannot be ruled out at this stage. Clinical benefit seems absent when administered after 1 week from onset of symptoms or in patients requiring ventilation, or in those who receive CCP with a low nAb titer. Nevertheless, chronically immunosuppressed patients benefit from CCP even at later stages (98, 105, 106) : the best evidence for this scenario comes from a prospective PSM showing a halving of mortality in ICU-admitted oncohematological COVID-19 patients who received CCP (107). We note that while there have been concerns that use in immuncompromised can promote the emergence of antibody-resistant variants, such variants have emerged from massive replication in susceptible populations and not from treated patients, who in any case are isolated in hospitals where mitigation efforts to reduce transmission are employed, and are thus very unlikely to transmit their viruses further (108). Such simple concepts have been poorly communicated to the general public and the clinical community, who should be better informed on the settings where CCP has shown efficacy and the ones where it has not.

### Recommendations

Stopping trials for futility is an occurrence that deserves special attention, because it represents wasted resources during a pandemic. Eight RCTs so far have been halted for futility, namely RECOVERY (49), REMAP-CAP (34), CONCOR-1 (109), C3PO (27), NCT04361253 (ESCAPE), CoV-Early (28), and COP20 (74), with the first one being to date the strongest evidence for futility (49), with its massive recruitment affecting the outcomes of systematic reviews (76). Instead of stopping trials for futility based on pre-set endpoints it makes more sense that DSMBs facing a high likelihood of lack of statistical significance provide advice on trial modifications that are likely to amplify the significance of signals of efficacy evident in these studies. This would seem a more responsible action than trial cessation given the paucity of therapeutic alternatives in the pandemic emergency.

More flexibility is needed when dealing with a new virus since estimates of efficacy and the required power can change tremendously as the data accumulates. For example, some RCTs were designed to evaluate efficacy at day 15 but subsequently we learned that this time was too early since mortality often occurs later (26). Indeed, a Bayesian re-analysis of RECOVERY data with a wide variety of priors (vague, optimistic, skeptical and pessimistic) calculated the posterior probability for both any benefit or a modest benefit (number needed to treat of 100). Across all patients, when analyzed with a vague prior, the likelihood of any benefit or a modest benefit was estimated to be 64% and 18% respectively. In contrast, in the seronegative subgroup, the likelihood of any benefit or a modest benefit was estimated to be 90% and 74% (79). This finding of benefit accruing to specific sub- groups, who were not determined post-hoc but because they were likely to benefit based on understanding of principles of CP treatment is found in nearly every trial whose overall finding is negative. This effect is more reflective of a problem with RCT design and execution than a limitation on the efficacy of CCP. Although we agree that subgroup analysis carries the risk of ‘cherry picking’ data, such analyses are often important for hypothesis generation and critically important during the emergency of a pandemic where neither viral pathogenesis nor therapeutic variables are well understood. When sub-group analyses are based on firm biological principles, such as focusing on those treated early in disease or lacking their own serological response, the exercise may be warranted. To emphasize this point, Christopher Columbus missed the pre-specified primary endpoint of his mission - reaching India - but no one considers his discovery of the New World to be a failure! Turning to the clinical arena, most trials of anticoagulants in myocardial infarction found reductions in mortality of about 20-25%, which was generally not significant in these underpowered trials that declared the findings to be null, even though such a mortality reduction would clearly be of value (110). Given that conventional peer-review slows down during a pandemic, pre-publishing RCT results by the preprint mechanism should be encouraged to accelerate sharing of potentially life-saving therapeutic approaches and to provide pre-publication review that could improve the quality of the final published study.

### The future of CCP

CCP remains a relatively inexpensive therapy that is available throughout the world even in resource poor areas that cannot afford expensive antiviral drugs or monoclonal antibody therapies. Much has been learned about the variables that affect CCP efficacy even though, as recounted here, the clinical efficacy data is mixed. Table 4 lists the RCTs whose outcomes have still to be reported after completion or which are still recruiting patients. Unfortunately, little new can be expected given that most of these RCTs were designed to enroll patients having symptoms for more than 7 days. Given the heterogeneity of the product and the complex variables that contribute to efficacy it is remarkable that many studies have reported reductions in mortality. This suggests a likely therapeutic effect that allow signals of efficacy to break through all the noise imposed by variability in the product and its clinical use. The positive evidence for CCP efficacy cannot be dismissed while in many cases negative results can be explained. In the absence of good therapeutic options for COVID-19, CCP is likely to find a niche in the early treatment of disease. Instead of looking for unlikely superiority outcomes, noninferiority RCTs comparing mAbs versus CCP in early arrivals should be initiated. Such an RCT is very unlikely to be sponsored by vendor companies, so public institutions should be sensitized to funding it.

There is evidence that vaccinated convalescents may have even higher nAb titers than unvaccinated convalescents (111), and that nAbs are more effective against VOCs than those from unvaccinated convalescents (112, 113), offering the promise of expanded success in using CCP. The higher frequency of high-titer donors would make donor screening more cost-effective. Being collected from individuals far from hospital discharge, and eventually periodic blood donors, it is unlikely that those units would benefit from further infectious risk minimization with PRTs, reducing the final cost and avoiding the previously discussed potential confounder of the inactivation method interfering with antibody function. Hence future CCP trials that will start after mass vaccination campaigns should preferentially rely over CCP from vaccinated donors. That said, inclusion of vaccinated individuals (e.g. CSSC-004 (29)) in future RCTs is also a potentially confounder. Indeed those individuals are at lower risk, thus increasing the outpatients NNTT to prevent a single disease progression, and also complicating the identification of seronegative individuals who are more likely to benefit from nAbs.

Low-to-middle income countries (LMIC) are likely to benefit the most from CCP, given they cannot afford massive deployment of mAb or small-chemical antivirals. Research is ongoing to spare some of cold chain requirements for CCP relying over freeze-dried plasma (FDP), which has been shown to preserve nAb functions for months at ambient temperature (114).

On November 8, 2021, the sudden emergence of the SARS-CoV-2 Omicron VOC with its 32 Spike mutations was an alarming development because it threatened to prolong the pandemic and undermine the immunity gains made with vaccination campaigns. Computer modeling and *in vitro* VNT soon confirmed full escape from clinically used mAb cocktails (including bamlanivimab+etesevimab, REGN-CoV-2 and AZD7442) (115, 116). While it was already known that mAb cocktails were less likely to experience immune escape than single mAbs, most of the scientific community underestimated the risk from major shifts in the 3D Spike structure. Combined with the lengthy manufacturing and approval process for mAb therapies, such sudden shifts largely hinder primarily mAb-based therapies, since polyclonal approaches such as CCP are less vulnerable to loosing activity. While even CCP is vulnerable to losing some efficacy with VOC that manifest partial immune escape, the probability and extend of reduction are far lower (117).

Given the experience accumulated with COVID-19, it is almost certain that CCP will again be considered to deal with the surges yet to come in the current epidemic as well as for the next epidemic. One of the good legacies of the COVID-19 pandemic is that an enormous international research effort had produced a large body of data that teaches how to best use CCP. The lessons learned reinforce the experience of the past, namely that best results are obtained with high titer CCP administered early in the course of disease. We are hopeful that lessons learned in this pandemic are heeded such that use and trials focus on the very early use with high-titer CCP.

We declare we have no conflict of interest to disclose.

## Data Availability

All data are available via request to the corresponding author

## Acknowledgements

this manuscript is written in memory of Dr.Giuseppe De Donno, Director of the Pneumology Division at the Mantua Hospital, a pioneer in the field of CCP.

## Author biographies

**Daniele Focosi**

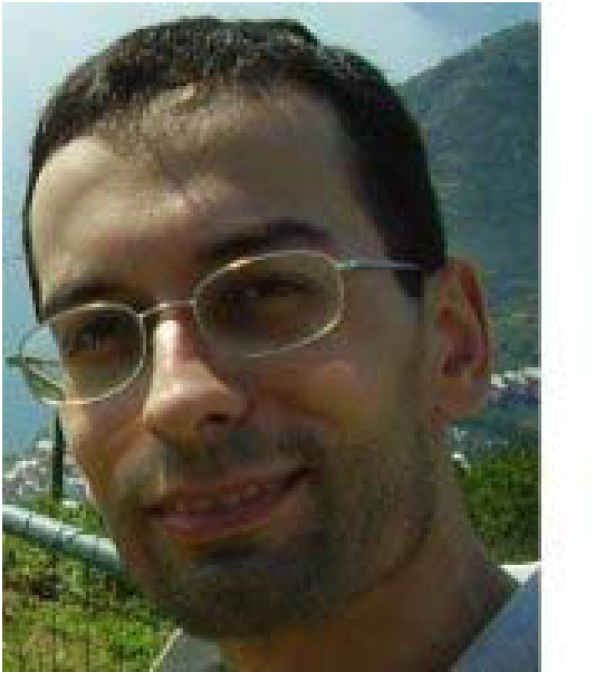

Dr. Focosi is a hematologist employed since 2009 as resident transfusion physician at the Pisa University Hospital. He has formerly been transplant immunologist and immunogeticist, quality assurance manager and production manager at the largest blood bank in Italy. He has received awards from the European Federation of Immunogenetics, the European Society of Organ Transplantation, and the Italian Society of Hematology. He has a Ph.D. degree in Clinical and Fundamental Virology, and a master degree in Clinical trials. He has authored 169 articles on topics ranging from emerging viral infections to new markers of immune compentence (including more than 50 and a SringerNature book on SARS-CoV-2 evolution), for a h-index of 30.

**Massimo Franchini**

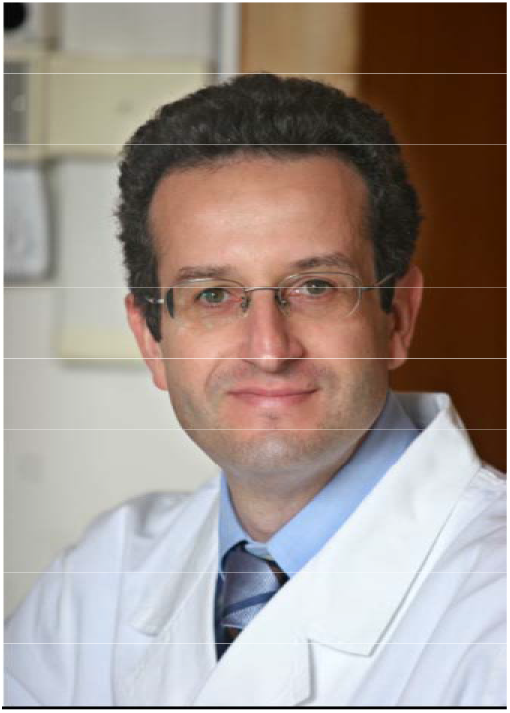

Dr. Franchini specialized in Hematology (1995) at the University of Verona (Italy). He is currently Director of the Department of Hematology and Transfusion Medicine of the Hospital of Mantua (Italy). He is associate Editor of Seminars in Thrombosis and Hemostasis, Member of the Committee of the Italian Association of Hemophilia Centers (AICE) for the revision of the Italian Guidelines on the management of Hemophilia. He is a member of the Regional Hematology Network (REL, Lombardy Region) - subcommittee of Hemostasis, and served as consultant of the Italian Ministry of Health – National Blood Center (2016–2020). His field of scientific research is predominantly dedicated to hemostasis and thrombosis. His is currently studying the COVID-19 convalescent plasma, from biological validation to its clinical use. He has authored more than 650 publications for a h-index of 83.

**Liise-anne Pirofski**

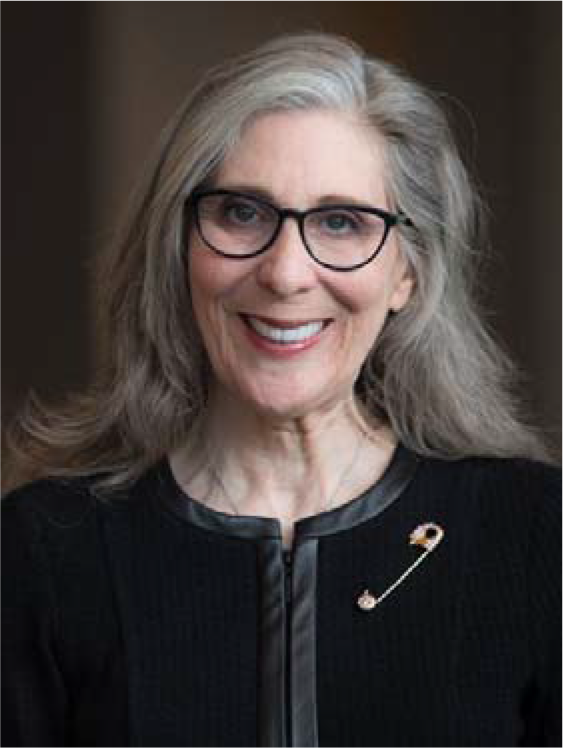

Dr. Liise-anne Pirofski is a physician-scientist. Her research programs focus on immunity to encapsulated pathogens and antibody mediated immunity to infectious diseases. She is chief of the Division of Infectious Diseases and the Jacques and Selma Mitrani Chair in Biomedical Research at Albert Einstein College of Medicine and Montefiore Medical Center. She is a member of the American Association of Physicians and a fellow of the American Academy of Microbiology, American College of Physicians, Infectious Diseases Society of America, and the American Association for the Advancement of Science. She is deeply devoted to biomedical education and mentoring for which she has received numerous accolades, including the American Society for Microbiology William A Hinton Award, and the Albert Einstein College of Medicine Faculty Mentoring Award, Harry Eagle Award for Outstanding Basic Science Teaching, and the Lifetime Achievement Award from the Albert Einstein College of Medicine Alumni Association.

**Thierry Burnouf**

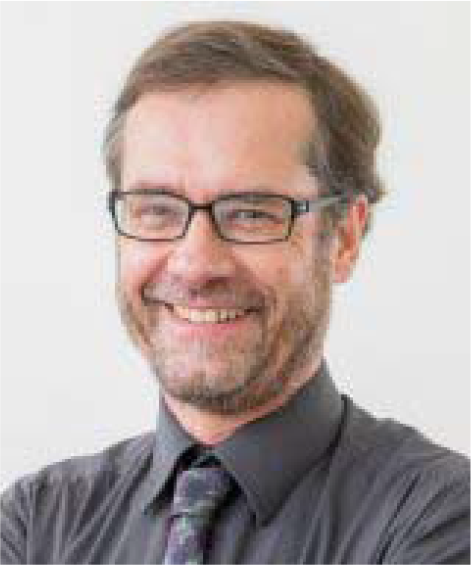

Dr. Burnouf is a protein biochemist who has authored more than 280 publications in the field of therapeutic blood products for a h-index of 53. He has received the 2019 Award from the International Plasma Fractionation Association. His current research interest focuses on Platelet growth factors for regenerative medicine and cell therapy, Plasma fractionation, Viral inactivation technologies, and Blood biotechnology. He is the Secretary of the Working Party on Global Blood Safety, and the Treasurer of the Working Party on Cellular Therapies of the International Society of Blood Transfusion.

**Nigel Paneth**

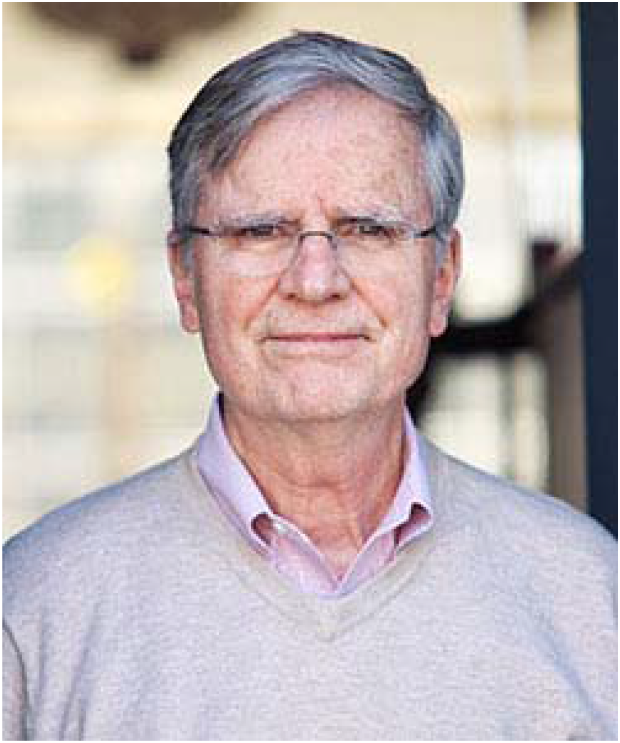

Dr.Paneth is a pediatrician and perinatal and child health epidemiologist with a particular interest in the causes and prevention of childhood neurodevelopmental handicap, especially cerebral palsy (CP). He was among the promotors of the US National COVID19 Convalescent Plasma Project (CCPP10). He is Emeritus University Distinguished Professor of Epidemiology and Biostatistics and Pediatricshas authored more than 320 publications for a h-index of 86.

**Michael J. Joyner**

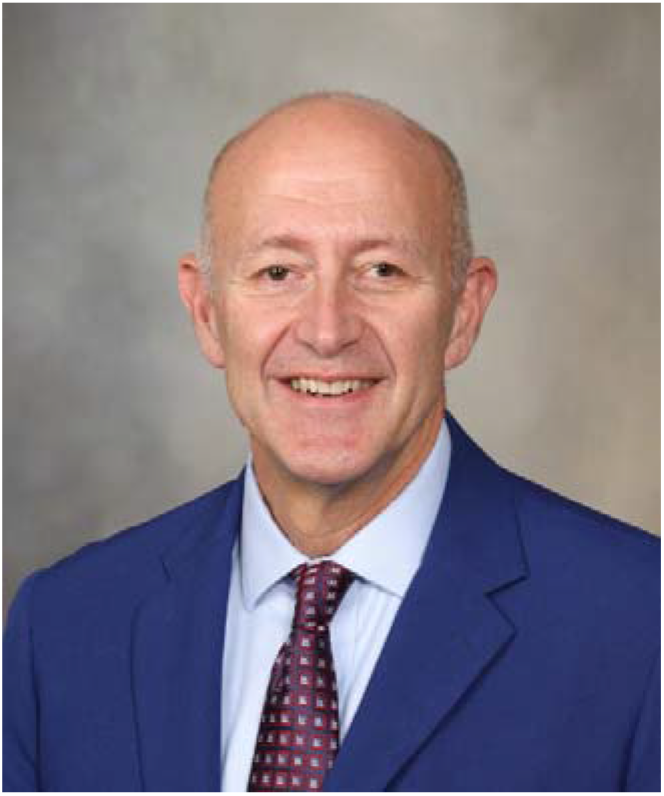

Dr.Joyner has authored more than 535 publications for a h-index of 93. His laboratory is interested in how humans respond to various forms of physical and mental stress during activities such as exercise, hypoxia, standing up and blood loss. Dr. Joyner is leading a national program sponsored by the U.S. Government to coordinate the collection and distribution of COVID-19 convalescent plasma for the treatment of individuals with severe or life-threatening disease.

**Arturo Casadevall**

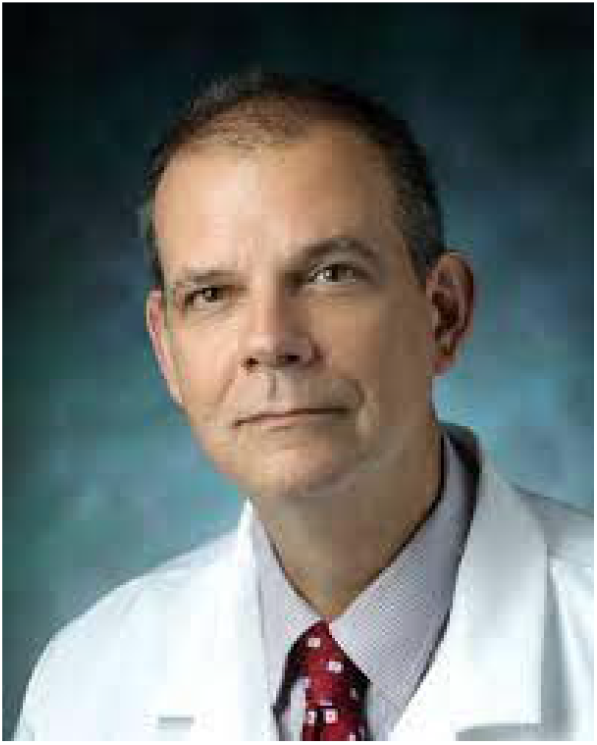

He is the chair of the Molecular Microbiology & Immunology, Bloomberg distinguished professor Alfred & Jill Sommer professor and chair professor. He has authored more than 900 PubMed-indexed articles for a h-index of 133. His research focuses on host defense mechanisms, how fungi cause disease, and in the development of antibody-based therapies for infectious diseases.

